# Escalating burden and mortality of carbapenem-resistant *Klebsiella pneumoniae* species complex infections in Bangladeshi infants

**DOI:** 10.1101/2025.06.11.25329357

**Authors:** Yogesh Hooda, Arif Mohammad Tanmoy, Naito Kanon, Hafizur Rahman, Mohammad Shahidul Islam, Zabed Bin Ahmed, Afroza Akter Tanni, Md Mobarok Hossain, Md Hasanuzzaman, Sharmistha Goswami, Tasnim Jabin, Rajib Chandra Das, Md Jamal Uddin, Belal Hossain, Shampa Saha, Anannya Barman Jui, Mohammod Shahidullah, Nobo Krishna Ghosh, AKM Shamsuzzaman, Nahid Sultana, Sanjoy Kanti Biswas, Feroza Aktar, Wazir Ahmed, Mohammed Monir Hossain, Mahbubul Hoque, ASM Nawshad Uddin Ahmed, Samir K Saha, Senjuti Saha

## Abstract

*Klebsiella pneumoniae* infections in young infants are an escalating threat in low- and middle-income countries, yet robust longitudinal data integrating hospital burden, clinical outcomes, antimicrobial, and genomics remains scarce. We performed an 18-year (2004-2021) prospective, multicenter genomic epidemiology study across four hospitals in Bangladesh. Among 122,353 enrolled children from whom blood or cerebrospinal fluid cultures were performed, 1,600 (1.3%) yielded culture-confirmed *Klebsiella pneumoniae* species complex (KpSC) isolates. Positivity increased from 16 per 1,000 cases tested in 2004 to 37 per 1,000 in 2021. Hospital case-fatality rate (CFR) rose from 21.4% to 51.4% during the study, paralleling the emergence and expansion of carbapenem resistance, first detected in 2008 and reaching 81% of isolates by 2021. Neonates accounted for 80.5% of infections and experienced a CFR of 40.8%. Whole-genome sequencing of 599 representative isolates revealed four KpSC species, 145 sequence types and 92 capsular alleles. Global high-risk clones ST11, ST16 and ST147 harbouring NDM-type carbapenemases dominated recent cases. These findings document the increasing resistance and mortality associated with KpSC infections amongst neonates in Bangladesh, underscoring the urgent need for strengthened infection prevention & control, equitable access to effective combination therapies, and vaccine-based preventative strategies.

## Introduction

Gram-negative bacteria of the genus *Klebsiella* are ubiquitous in the environment and form part of the commensal flora of healthy humans^1,2^. Among them, *Klebsiella pneumoniae* and six related members of the *K. pneumoniae* species complex (KpSC) can also cause a broad spectrum of infections, including soft tissue, surgical wound, urinary tract, respiratory tract, bloodstream and central nervous system infections^3^, particularly in hospital settings and immunocompromised individuals. Neonates (0-27 days old) are particularly susceptible to systemic infections by KpSC^4–8^.

Reports of multidrug-resistant (MDR) *K. pneumoniae* have risen sharply worldwide over the past two decades, with the steepest increases observed in low and middle income countries (LMICs)^9–11^. The emergence of carbapenem-resistant *K. pneumoniae* is associated with high mortality rates^12^, and has further constricted therapeutic options, leading to it being declared a critical priority pathogen by the WHO.

With the antibiotic pipeline faltering, alternative strategies are gaining traction. Recently, combination drugs that contain a beta-lactam antibiotic and a beta-lactamase inhibitor have shown to be efficacious for treatment of carbapenem resistant *K. pneumoniae* infections, but their activity depends on the underlying carbapenemase gene as these do not work on all family of metallo-β-lactamases.^13^ Passive immunotherapies and maternal vaccines targeting capsular (K) or lipopolysaccharide (O) antigens are also under development^14–17^. However, *K. pneumoniae* exhibits extraordinary antigenic heterogeneity, with at least 13 O-types and 186 K-types documented to date^18^. Selecting vaccine valences therefore demands precise, contemporary data on circulating serotypes, resistance genes and clonal backgrounds.

Additionally, global genomic sequencing efforts have found that other KpSC members such as *K. quasipneumoniae* and *K. variicola* cause invasive diseases in humans, yet routine biochemical tests cannot differentiate them from *K. pneumoniae*^2^. Failing to recognize this hidden diversity can mask transmission chains, underestimate true burden and misguide therapy, particularly in low-resource settings that rely on phenotypic methods.

In this study we: (i) quantify temporal trends in the hospital burden and severity of invasive KpSC infections in Bangladeshi children; (ii) describe the evolution of phenotypic and genotypic AMR, including carbapenemase rise and spread; and (iii) characterize the species, sequence types, O- and K-loci of a representative subset of isolates using whole-genome sequencing. Overall, we generate data to inform future infection-control measures, empirical-therapy guidelines and multivalent-vaccine design in Bangladesh and neighboring countries.

## Results

### The burden and severity of KpSC infections in Bangladesh is increasing

Between 2004 and 2021, 122,353 cases had blood and/or CSF samples cultured at the treating physician’s discretion (Sup Fig. 1); all were enrolled in this study. Bacteriological-culture and biochemical testing confirmed that 1,600 (1.3%) infections were caused by the KpSC. A sharp rise is observed in the number of KpSC isolates starting in 2018 (Fig. 1a). Up to 2017, the mean positivity rate for KpSC was 16 per 1,000 cases where blood cultures were performed, which climbed to 18, 34 and 37 per 1,000 suspected sepsis cases in 2019, 2020 and 2021, respectively (Fig. 1b). Seventy-four percent (1,182/1,600) of *KpSC-*positive cases were reported from one hospital, BSHI (Sup Fig. 2a), which also accounted for the highest positivity rate among the four hospitals since 2015 (Sup Fig. 2b).

**Figure 1:**
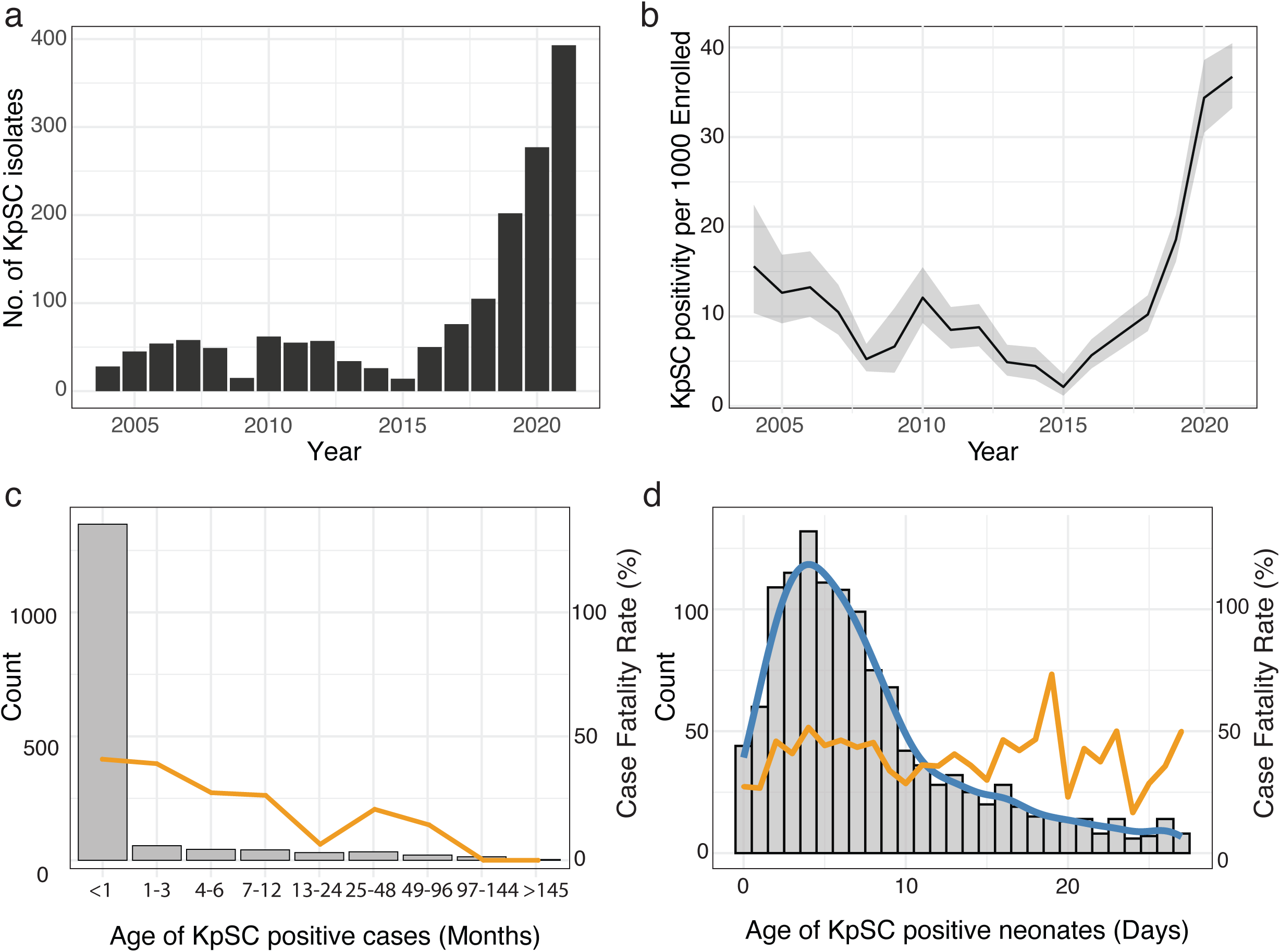
Number and age distribution of culture-confirmed KpSC infections. **a,** Annual number of KpSC isolates, 2004 – 2021. **b,** Annual positivity rate (per 1,000 blood cultures performed). **c,** Age distribution of all KpSC-positive cases (bars, grey) with case fatality rate by age in months (line, yellow). **d,** Age distribution of KpSC-positive neonates (total numbers in bars, grey; and distribution of neonates in line, blue) with case fatality rate in days (line, yellow).

The median age of KpSC-positive cases at hospital admission was 5 days (IQR: 2-18) and at blood draw was 8 days (IQR:4-21) (Fig. 1c). Overall, 1,352/1,600 (84.5%) KpSC infections occurred in infants <2 months old, and 1,288/1,600 (80.5%) in neonates (first 28 days old). The hospital case fatality rate (CFR) was highest during the first month of life (41.5%) and declined for infants older than three months (Fig. 1c). Overall CFR was 38.1%. Among neonates, the median age at sample collection was 6 days (IQR: 3-21), and the CFR was consistent throughout the first month of life (Fig. 1d). Most infections 1,076/1,600 (67.3%) were detected within 72 hours of admission (Sup Fig. 3a); the median interval between admission to sample collection was 24 hours (IQR: 0-4).

The severity of KpSC infection is highlighted by hospital outcomes: 610/1,600 (38.1%) cases died at the hospital, 174/1,600 (10.9%) left against medical advice, and 816/1,600 (51.0%) were discharged from the hospital (Sup Table 1). The average hospital stay was 8 days (IQR, 4-14), but 64% babies who died, succumbed within the first 5 days of admission (Sup Fig. 3b). CFR started increasing in 2016 (Fig. 2a), with the odds of death in 2021 being 3.9 times higher than in 2004 (95 % CI 1.5–9.8; Fig. 2b). The proportion of patients leaving hospital against medical advice varied over the study period, ranging from ∼4% to 20% per year, without a consistent increasing trend over time (Sup Fig. 4a). Sensitivity analyses incorporating alternative assumptions regarding LAMA outcomes demonstrated that the temporal increase in mortality was preserved across all scenarios (Sup Fig. 4b).

**Figure 2:**
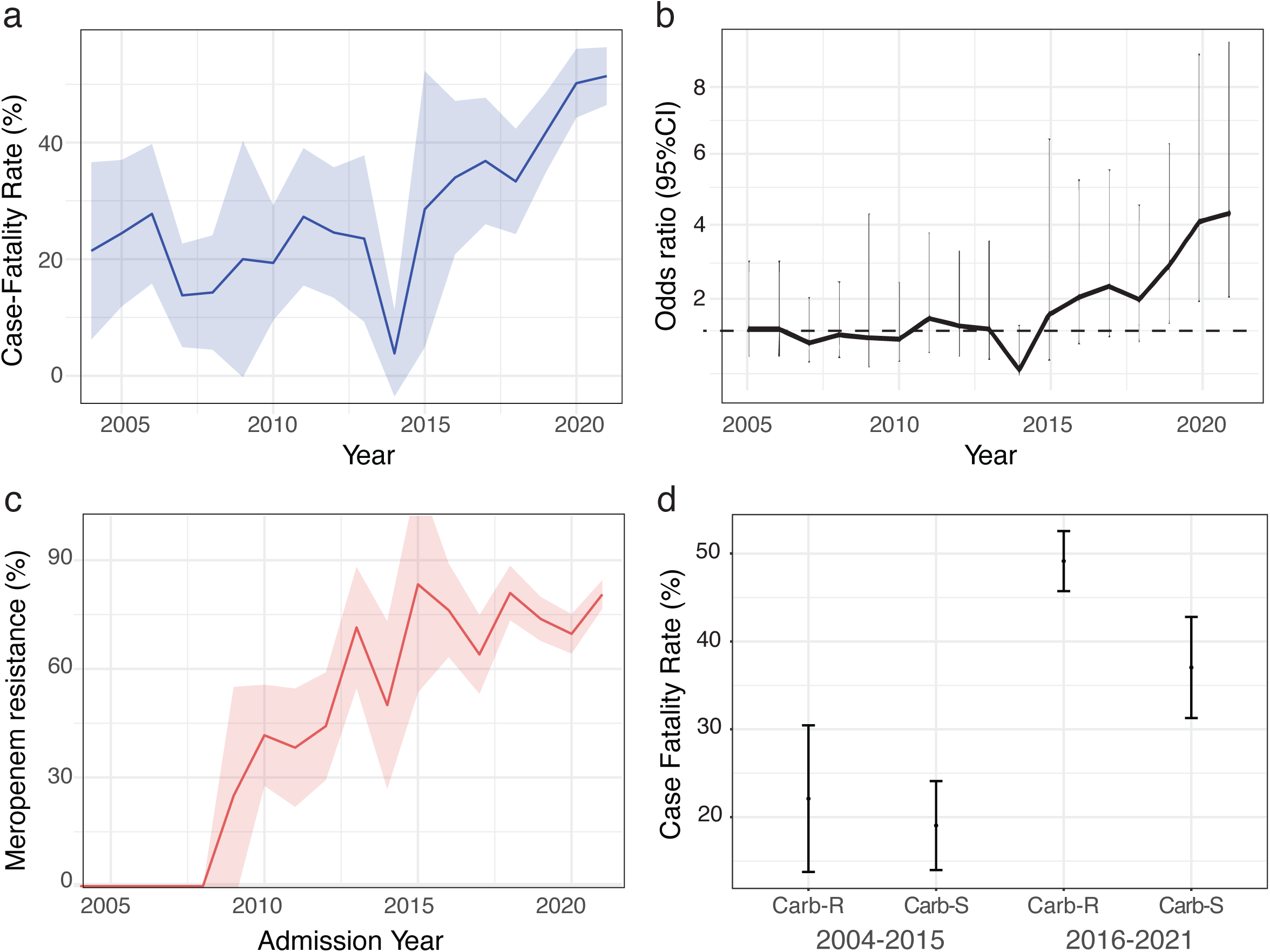
Rising case fatality rate of KpSC infections and correlation with rising carbapenem resistance. **a,** Yearly trend in case-fatality rate (in %) amongst KpSC cases at the study hospitals. **b,** Odds ratio (95%CI) of mortality amongst KpSC positive cases using 2004 as the base year. **c,** The yearly trends in the percentage of KpSC isolates that were resistant to meropenem. **d,** The comparison of case-fatality rate (in %) of meropenem-resistant and meropenem-sensitive KpSC isolates stratified for: 2004-2015 and 2016-2021.

### Increasing severity of KpSC infections correlates with rise in carbapenem resistance

We investigated the possible drivers of escalating mortality by comparing yearly trends in clinical features (Sup Fig. 5) and antimicrobial susceptibility profiles for 10 antibiotics (meropenem, imipenem, ceftriaxone, ampicillin, gentamicin, netilmicin, ciprofloxacin, amikacin, chloramphenicol, and cotrimoxazole) proposed by CLSI^19^ (Sup Fig. 6). KpSC isolates from Bangladesh exhibited high resistance against most antimicrobials used, including amikacin, cotrimoxazole, gentamicin, netilmicin, and ciprofloxacin. The first case of carbapenem resistance was detected in 2008, and with noticeable increase during the study period, by 2021 >80% of KpSC isolates were resistant to carbapenem (Fig. 2c). The rising carbapenem resistance correlated with increasing mortality, with the odds of death being 3.2-fold higher (95% CI: 2.3-4.6) relative to carbapenem susceptible isolates (Sup Fig. 7).

To further assess the impact of carbapenem resistance, we stratified cases into two epochs, 2004–2015 and 2016–2021, and compared CFRs for carbapenem-resistant versus carbapenem-susceptible KpSC infections (Fig. 3d). During 2004–2015, CFRs were similar irrespective of resistance phenotype. From 2016 onward, carbapenem-resistant infections exhibited a markedly higher CFR than susceptible infections, however, mortality increased for both groups. In multivariable logistic regression analyses, carbapenem resistance remained consistently associated with increased odds of in-hospital CFR across all models, including those with individual adjustments for covariates of age group, sex, hospital site, timing of culture collection, and admission epoch (Sup Fig. 8). Upon simultaneous adjusting for neonatal status, hospital site, and admission epoch, the estimated effect size for carbapenem resistance was attenuated. This likely reflects shared variance between carbapenem resistance, temporal trends, and referral patterns, as well as reduced effective sample size, rather than absence of an association. Taken together, these analyses suggest, while carbapenem resistance is a key driver of poor outcomes, additional factors, i.e. host, pathogen and/or environmental, have contributed to the post-2016 surge in KpSC-associated deaths.

**Figure 3:**
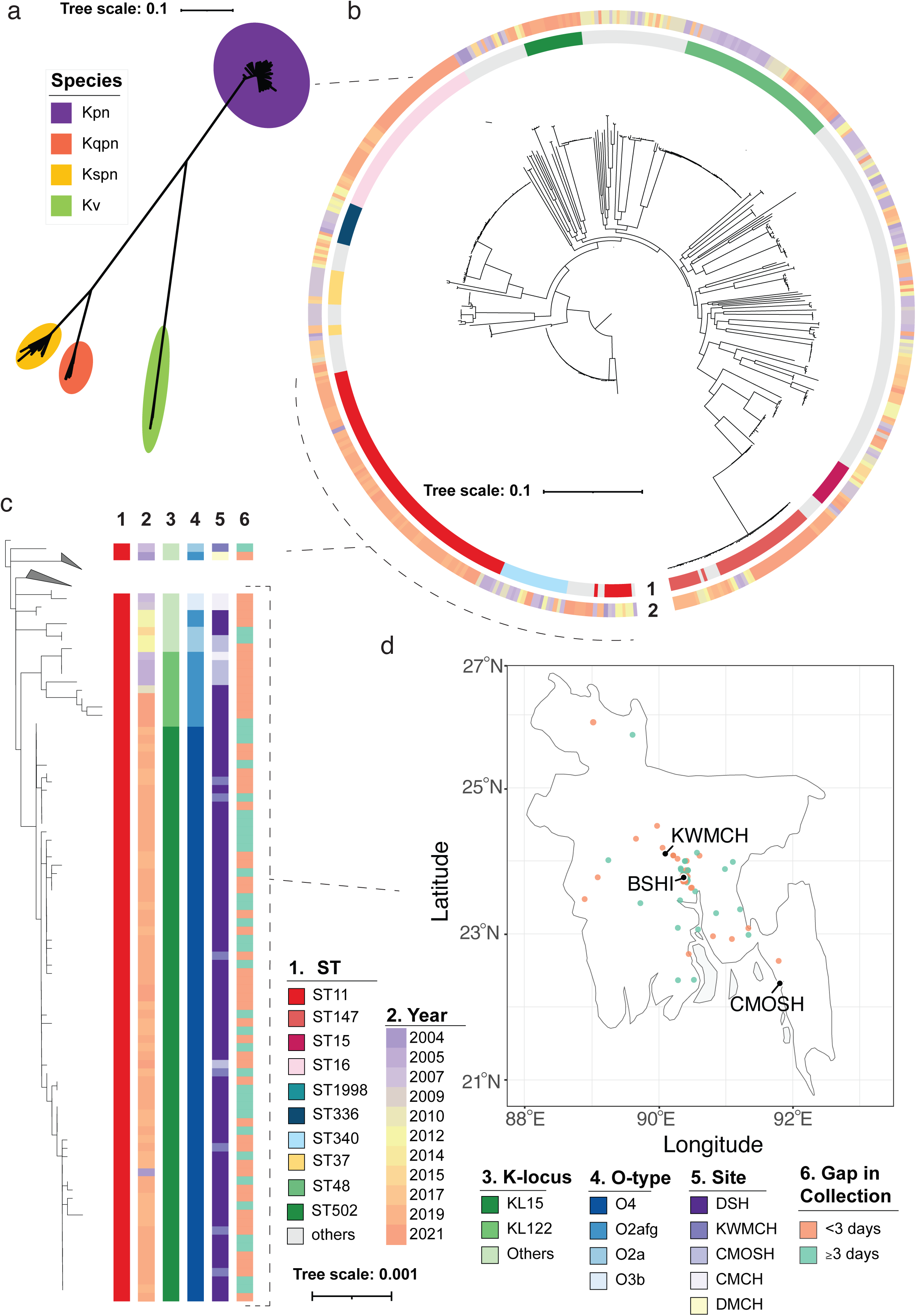
Phylogenetic structure and temporal dynamics of *Klebsiella pneumoniae* species complex isolates. **a,** Maximum likelihood phylogenetic tree of 599 representative KpSC genomes, colored by species. Kpn: *K. pneumoniae*, Kqpn: *K. quasipneumoniae subsp quasipneumoniae*, Kspn: *K. quasipneumoniae subsp similipneumoniae*, Kv: *K. variicola*. **b,** Maximum likelihood phylogenetic tree of the 503 *K. pneumoniae* genomes, annotated with year of isolation and sequence type (ST). **c,** Maximum-likelihood tree showing the genomes belonging to ST-11 highlighting a large cluster of *K. pneumoniae* ST-11 in 2019-2021. The accompanying heatmap summarizes epidemiological variables (year of isolation, hospital site, interval between admission and blood draw) and genomic features (ST, O locus, K type) for the ST11 genomes. **d,** Map of Bangladesh showing the home location of patients involved in the *K. pneumoniae* ST11 cluster who presented at the study hospitals. Points are colored by the interval between admission and blood draw, indicating a country-wide cluster.

### KpSC infections exhibit high genomic and antigenic diversity

To further characterize the KpSC isolates, we sequenced a representative subset of 527 isolates (527/1461, 36.1%) collected across the entire study period from infants younger than two months - the age group with the highest burden (Sup Fig. 9a, Sup Table 2, Sup Data 1)^26^. Some bio banked strains from 2004-2009 could not be revived, and hence fewer representatives from those years were available. To partially overcome this limitation, we included 72 sequences from three other hospitals from 2004-2009 (Sup Table 2, Sup Data 1). Among the total 599 isolates sequenced, 503 (84.0%) genomes were *K. pneumoniae*, 72 (12.0%) were *K. quasipneumoniae* subsp *similipneumoniae*, 20 (3.3%) were *K. quasipneumoniae* subsp *quasipneumoniae*, and 4 (0.7%) were *K. variicola* (Fig. 3a). The annual distribution of these species remained broadly consistent throughout the study period (Sup Fig. 9b). Infections caused by *K. pneumoniae* were associated with a 2.18 (95% CI 1.06-4.94) fold higher case fatality rate than infections due to other KpSC species (Sup Fig. 10).

Maximum-likelihood phylogenies of the 599 sequenced KpSC genomes (Fig. 3a) and the 503 *K. pneumoniae* genomes (Fig. 3b) demonstrated high genetic diversity amongst KpSC isolates identified in this study. We identified 145 distinct sequence types (STs); however, the ten most common STs accounted for 52% of all isolates. The predominant lineage was *K. pneumoniae* ST11, 97% of which were identified between 2019 and 2021. These ST11 genomes were sampled across three of the four surveillance hospitals in Dhaka, Chattogram and Mirzapur, yet showed minimal within-lineage diversity, consistent with a nationwide clonal expansion (Fig. 3c, Sup Fig 11). Metadata reinforcing this observation include that among ST11 cases, the median interval from admission to blood draw was 2 days (IQR 0-5); 53% (36/68) of samples were collected within 48 hrs of hospital admission (Fig 3c). Mapping of home addresses shows infants originating from 22 districts (Fig. 3d).

The yearly distribution of the 10 most common STs for *K. pneumoniae* are shown in Fig. 4a. Some of the common STs like ST11, ST147 and ST16 exhibited association with higher CFR (Sup Fig. 10), however when adjusted for carbapenem resistance, barring ST11, no other ST exhibited statistically significant association with higher mortality (Sup Table 3). Compared to a global database of *K. pneumoniae,* most of the common STs in Bangladesh have been reported in other countries, and ST11, 16, and 147 belong to global problem clones (Fig. 4b).

**Figure 4:**
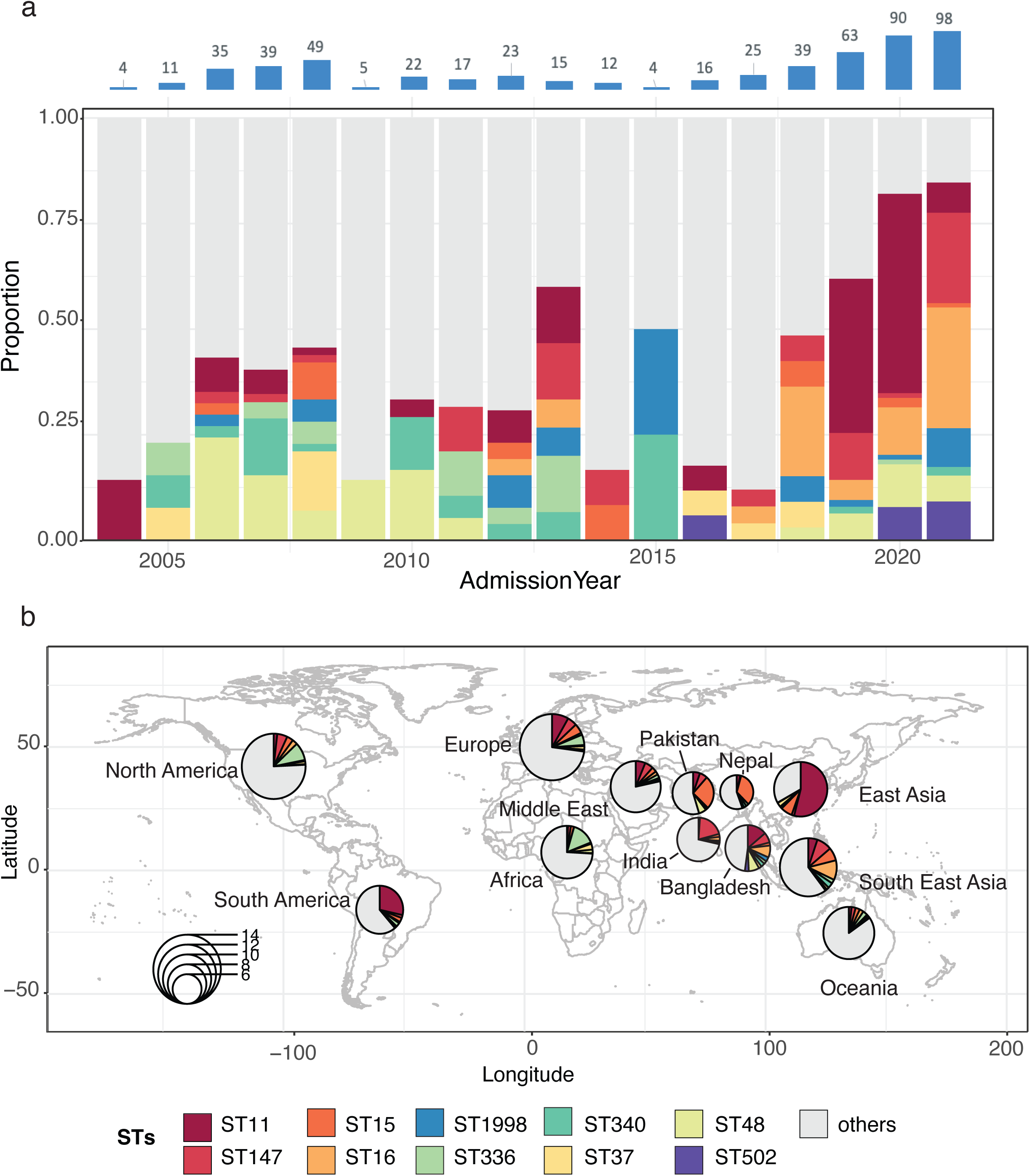
Temporal distribution of dominant *Klebsiella pneumoniae* sequence types in Bangladesh and their global prevalence. **a,** Annual proportions of the ten most common sequence types (STs) detected in Bangladesh, 2004–2021; colors correspond to individual STs. **b,** Prevalence of the ten STs among a global collection of 32,656 *K. pneumoniae* genomes. Red outlines denote the three recognized global problem-clone lineages (ST11, ST16, ST147).

We next analyzed genomic carbapenem-resistance and virulence determinants in the sequenced KpSC genomes and compared these with meropenem susceptibility results, and mortality respectively. Genomic prediction of the meropenem phenotype achieved a sensitivity of 96.3% and a specificity of 65.4% and matched the phenotypic carbapenem trendline (Sup Fig. 9c). The main carbapenemase alleles detected were blaNDM-5, blaNDM-1, along with multiple other less common variants (Table 1), indicating that different KpSC lineages in Bangladesh have acquired carbapenem-resistance genes independently. For virulence, we specifically looked for presence of genes encoding siderophore-uptake systems (aerobactin and salmochelin) and hypermucoid phenotype (RmpA/RmpA2) (Sup Fig. 10). None of the known virulence factors were associated with increased mortality, as none of the dominant carbapenem-resistant strains (ST11, ST16 and ST147) contained these virulence factors. In 2019, we identified a single ST43 isolate with both bla-OXA181 and aerobactin encoding genes. While this was an isolated case, such carbapenem-resistant hypervirulent strains should be continuously reported.

**Table 1:** List of carbapenem-resistance genes present on 599 KpSC genomes in this study.

| **Resistance marker** | **STs present** | **Frequency** | **First documented** |
| --- | --- | --- | --- |
| blaNDM-5 | 11, 147, 48, and 7 others | 126 | 2016 |
| blaNDM-1 | 502, 16, 307 and 35 others | 113 | 2008 |
| blaNDM-5,blaOXA-181 | 16 and 147 | 38 | 2018 |
| blaNDM-7 | 1998, 11, 705 and 6 others | 21 | 2012 |
| blaNDM-5;blaOXA-232 | 147, 307 and 437 | 14 | 2019 |
| blaKPC-2;blaNDM-1 | 1998, 231, 340 and 70 | 12 | 2015 |
| blaOXA-181 | 14, 16 and 43 | 4 | 2012 |
| blaKPC-2 | 1998, 273 and 334 | 3 | 2019 |
| blaNDM-1;blaOXA-232 | 14 | 3 | 2016 |
| blaNDM-4 | 1584-1LV | 2 | 2019 |
| ~blaNDM-5 | 152 | 2 | 2019 |
| blaOXA-232 | 231 | 2 | 2018 |
| blaNDM-24; blaOXA-181 | 16 | 1 | 2018 |
| blaNDM-5; blaOXA-48 | 437 | 1 | 2021 |

To characterize antigenic diversity relevant to vaccine design, we analyzed the genomic regions encoding the capsular polysaccharide (K locus) and O antigen. In total, 92 unique K loci and 11 unique O loci were identified. The ten most prevalent O loci and K loci together accounted for 99.8% (Fig. 5a, Sup Fig. 12) and 58.3% (Fig. 5c, Sup Fig. 13) of all isolates, respectively, with year-to-year variations (Fig. 5b,5d). The combined share of the ten dominant O loci stayed above 95% every year, whereas the share of the ten dominant K loci was highly variable, oscillating between 0% and 86.7%.

**Figure 5:**
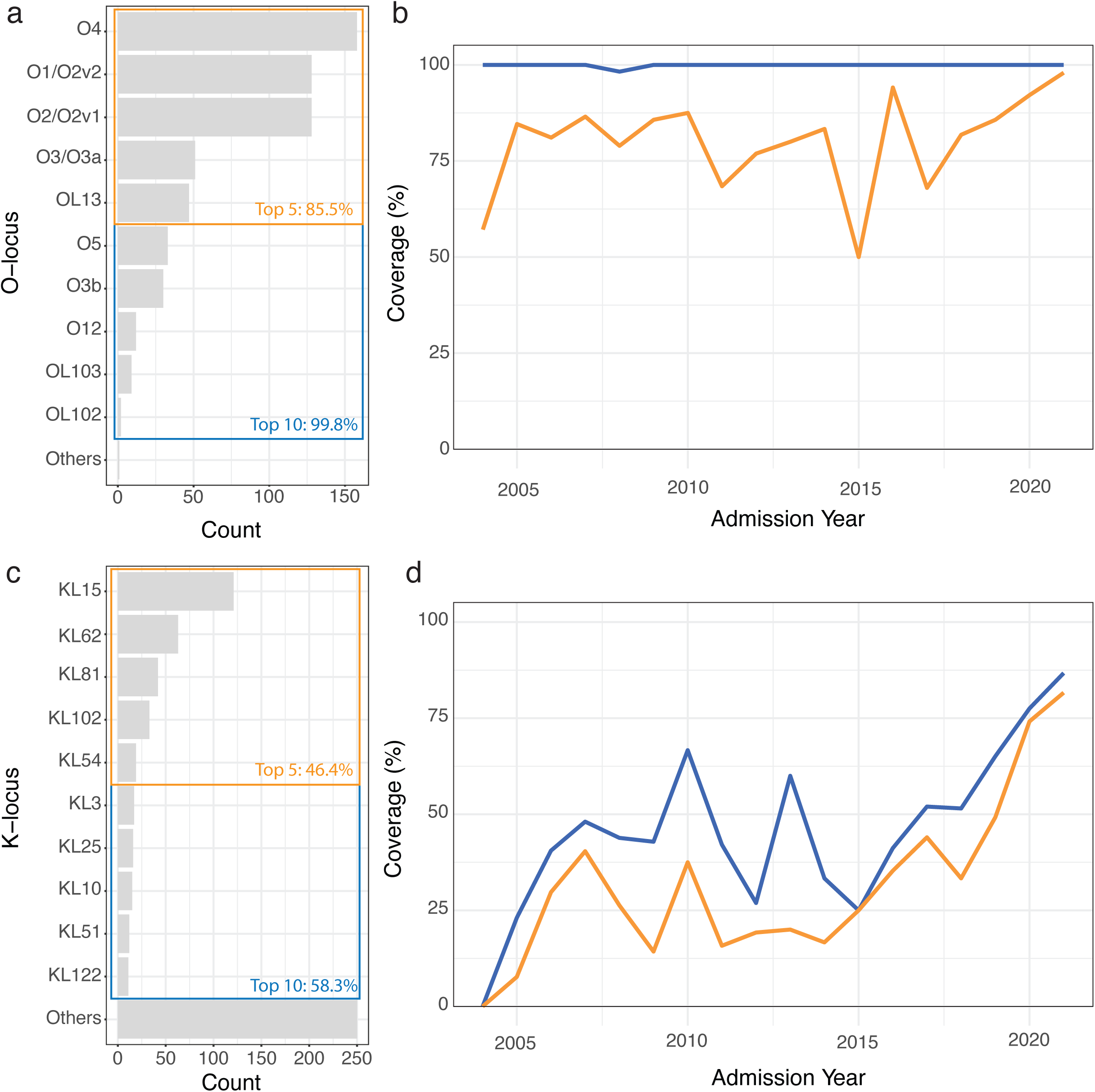
Frequency and temporal coverage of dominant O- and K-loci among the KpSC isolates. **a,** Absolute counts of the ten most common O-loci. Colored boxes denote the cumulative coverage provided by the five (orange) and ten (blue) leading O-loci. **b,** Annual percentage coverage achieved by the five (orange) and ten (blue) most common O-loci. **c,** Absolute counts of the ten most common K-loci. Colored boxes denote the cumulative coverage provided by the five (orange) and ten (blue) leading K-loci. **d,** Annual percentage coverage achieved by the five (orange) and ten (blue) most common K loci.

## Discussion

Geographically and temporally representative epidemiological and genomic data from high-burden LMICs are essential to guide policy and intervention against KpSC infections. Here we report 18 years of surveillance from four Bangladeshi pediatric hospitals, detailing trends in burden, antimicrobial resistance and genomic diversity.

From 122,353 bacteriological culture of blood and/or CSF samples conducted between 2004 and 2021, we identified 1,600 KpSC isolates. Neonates comprised 80.5% of cases and had a hospital case-fatality rate of 40.8%, compared with an overall CFR of 38.1%. In a multi-country study on neonatal infections, BARNARDS, conducted in seven LMICs including Bangladesh from Nov 2015 to Dec 2017, the CFR due to KpSC infection was 22.4% (58/258), compared to 35.7% (45/126) during the same period in this study.^6^ In the NeoObs study done across 11 countries from 2018-2019 including Bangladesh, neonatal KpSC case fatality rate was 21.1%.^7^ In NeoOBS Bangladesh site, the CFR was 22% (2/9), compared to 39% (120/307) during that period in this study. Within neonates, infections were most commonly detected on day 5, as seen in previous studies [BARNARDS and NeoObs]^7,8^. Collectively, these studies highlight the burden of KpSC infections amongst neonates globally. The higher mortality we observed was driven by BSHI, which functions as the pediatric referral center for Bangladesh, receiving the most critically ill infants from across the country; this referral bias is likely to inflate both the apparent burden, and the CFR compared with primary or secondary facilities included in the other studies.

Overall, the alarming rise in neonatal deaths threatens to erode vaccine-driven gains in child survival across LMICs. Thirteen percent of infections were detected within the first three days of life, including 3% within 24 hours of birth, and 67% of infants were culture-positive within 72 hours of admission. These timelines point to transmission in maternity units, delivery clinics, or the community, prior to referral to the study hospitals.

At BSHI, the KpSC positivity rate rose from 16 per 1000 suspected sepsis cases in 2004 to 37 per 1000 in 2021, and the hospital-specific case-fatality rate climbed from a mean of 20.7% (2004–2014) to 51.4% in 2021. Carbapenem resistance was associated with death, with an adjusted odds ratio of 3.2 (95% CI: 2.3-4.6). A positive correlation between carbapenem resistance and mortality has reported previously for KpSC and other bacterial infections^34^; however, the observational design of our study precludes inferring causality. Consistent with this association, the case-fatality rate of carbapenem-resistant KpSC infections was significantly higher than that of carbapenem-sensitive KpSC infections (46.3% vs 25.4%). Notably, mortality among carbapenem-susceptible infections also increased after 2015 compared with earlier (31.1% vs 20.3%). Adjustment for neonatal status, admission time, and hospital site reduced precision of carbapenem resistance mortality effect estimates. These factors are likely themselves closely linked to the emergence and clinical impact of carbapenem-resistant KpSC and thus may lie on the causal pathway rather than acting as independent confounders.

The excess mortality in carbapenem-susceptible cases after 2015 suggests that factors beyond drug resistance are driving severity. Several, not mutually exclusive, hypotheses warrant exploration: (i) bacterial adaptation – dominant lineages may have acquired additional virulence determinants^3^ or stress-tolerance genes that enhance survival in neonatal wards^35^; (ii) changing obstetric practice – the sharp rise in Caesarean deliveries in Bangladesh increases early hospital exposure and separates infants from protective maternal microbiota^36,37^; (iii) ward overcrowding and resource strain – more institutional births and limited isolation facilities hamper effective infection-prevention and control^38^; (iv) referral bias – improved road networks now funnel the sickest infants who fail first-line therapy or need advanced support to BSHI, inflating observed mortality^39^ (v) comorbidities and other host-factors **–** rising prevalence of prematurity, low birth-weight and congenital anomalies may increase vulnerability to sepsis^40–42^; and (vi) climate stress – warmer, more humid conditions, extreme weather events like flooding and heat-related infrastructure strain can favor environmental persistence and spread of KpSC within hospitals^43,44^. These possibilities highlight the need for integrated clinical, microbiological and health-systems investigations to disentangle drivers of the continued rise in KpSC-associated neonatal deaths.

Several genomic studies have now established that biochemically identified *K. pneumoniae* infections can be other members of KpSC^2,26,33^. In this study, 15% were *K. quasipneumoniae* (*subsp. simillipneumoniae* & *subsp. quasipneumoniae*) and *K. variicola*, rather than *K. pneumoniae*. Infections due to *K. pneumoniae* carried a higher hospital CFR (32.9% vs 18.3%) and a greater burden of carbapenem resistance genes. These observations suggest that pharmacological interventions directed solely at *K. pneumoniae*, whether vaccines, monoclonal antibodies or new antimicrobials, could create ecological space for the related species to fill. Future control strategies must therefore target the entire KpSC repertoire to avoid pathogen replacement and should also include broad IPC.

We also observed a high diversity of carbapenem-resistance determinants in Bangladeshi KpSC isolates, with the *blaNDM-1* gene predominating^6,45^. Because KpSC isolates are predicted to remain susceptible to the ceftazidime-avibactam + aztreonam combination ^13^, this regimen is a promising therapeutic option. Yet neither drug is widely available in Bangladesh due to their cost, particularly in the low-income communities served by our surveillance hospitals, creating a critical treatment gap which needs to be addressed through changes in drug procurement recommendations/policies. Clinical trials are therefore urgently needed in Bangladesh to evaluate the regimen’s efficacy, safety and implementation feasibility and to guide policy aimed at reducing KpSC-associated neonatal mortality.

Overall, the Bangladeshi KpSC population is highly heterogeneous: the 599 genomes were distributed across 145 STs, and the dominant lineages shift and vary over time. Notably, several dominant sequence types that emerged or expanded after 2015 accounted for a large fraction of deaths, suggesting that clonal shifts may have contributed to the observed increase in case fatality. Notably ST11, ST147 and ST16 have been isolated in various other countries^26,33^. The clinical impact is best illustrated by ST11, that is associated with 2.41 times (95% CI:1.39–4.24, p = 0.001) increased odds of mortality compared to other KpSC strains, even after adjusting for carbapenem resistance. Almost all ST11 isolates (n = 68) were recovered between 2019 and 2021, consistent with a recent clonal expansion. None of the ST11 isolated possessed any known virulence factors such as aerobactin, salmochelin and RmpA. The median interval from admission to blood draw was 24 hours (IQR 0–5); 53% of cultures were taken within 72 h of hospital admission. Cases originated from 22 districts, and most infants were admitted directly from home or local clinics rather than transferred from other hospitals. Minimal genetic divergence, very short admission-to-culture intervals and wide geographic scatter together suggest transmission in maternity units, delivery clinics or the community, settings upstream of tertiary infection-control measures. Future studies to identify host, exposure, and care-related risk factors associated with infection by high-risk lineages and poor outcomes are urgently needed. Such integrated approaches will be essential to inform interventions that are tailored to the evolving clonal structure of KpSC in hospital settings, especially neonatal wards.

Efforts to protect young infants against KpSC, particularly *K. pneumoniae*, now focus on maternal vaccines and monoclonal antibodies that target the organism’s surface polysaccharides (capsular K antigen and lipopolysaccharide O antigen)^16^. Our data highlight the antigenic hurdles such products must clear. Whole-genome analysis predicted 92 distinct K (capsular) types, with the ten most common covering only 58% of isolates. O-antigen diversity was narrower: the ten most frequent O loci encompassed 99% of isolates, and their cumulative coverage remained >50 % each year. Antibody access to the O-antigen may be partially constrained by the thick polysaccharide capsule, which can shield the underlying lipopolysaccharide and limit immune recognition in heavily encapsulated strains^46^. By contrast, K-type coverage fluctuated markedly over time, reflecting turnover of dominant sequence types and the strong linkage between ST and capsule genotype. These patterns indicate that a multivalent formulation combining O- and K-antigens will be required for broad protection in Bangladesh, yet the capsule’s extensive and shifting diversity will remain a major design challenge.

The findings of this study should be viewed within the context of certain limitations. First, we did not collect information related to gestational age, birth weight and maternal factors, such as maternal age, nutrition, referral status (inborn vs outborn), standardized measures of illness severity at admission (e.g. SNAPPE-II) and mode of delivery. These variables are recognized risk factors for KpSC infection, and their absence prevents adjustment for host susceptibility. Second, our study lacked systematic data on comorbidities including congenital anomalies, which may have contributed to the observed mortality; nevertheless, previous CHAMPS investigations have implicated *K. pneumoniae* as a leading cause of neonatal death.^8^ Third, no pre-hospital records were available, so we could not ascertain prior antibiotic exposure, treatment adequacy or the likely place of acquisition,, illness severity at admission, hospital bed occupancy, staffing levels, and time to antibiotic administration. These factors may plausibly contribute to rising mortality andthe lack of such data limits our ability to disentangle pathogen-specific effects from broader system-level drivers of mortality. Fourth, we sequenced a representative subset of KpSC isolates; rare sequence types, capsular types or O-antigen variants present in the isolates that were not sequenced may therefore have been missed. Fifth, automated blood culture system (BACTEC) was introduced in BSHI, KWMCH and MRKSH in 2016 and in CMOSH in 2017. Because parallel manual and automated cultures were not performed during the transition, we lack internal cross-validation data, precluding calculation of a reliable adjustment factor to account for this methodological change. Although automation is likely to improve overall pathogen recovery and reduce time to positivity, the rise in KpSC burden and mortality was not temporally synchronous across sites, was driven predominantly by BSHI, continued to increase for several years after implementation rather than showing an abrupt step change, and was selectively enriched for carbapenem resistant lineages. Moreover, prior studies suggest that automated systems increase overall positivity primarily through improved detection of fastidious organisms, with similar recovery of common pathogens such as *K. pneumoniae* across methods^47^. Taken together, BACTEC is unlikely to explain the magnitude and selective nature of the observed increase. Despite these constraints, the dataset provides one of the most comprehensive clinical–genomic reports of paediatric KpSC infections available from South Asia. Rising incidence, a four-fold increase in mortality and the dominance of blaNDM-positive global clones expose a critical threat to neonatal survival. To mitigate this risk, policies should include: (i) expansion of neonatal intensive-care capacity and skilled staffing at birth facilities; (ii) stringent infection-prevention and control in labour wards and neonatal units; (iii) continuous, denominator-based genomic surveillance to detect clonal shifts and emerging resistance; and (iv) rapid access to effective therapeutics, including context-specific trials of ceftazidime–avibactam plus aztreonam. Maternal vaccines or monoclonal antibodies targeting the prevalent O-antigens could add a valuable layer of protection, but the extensive and shifting capsule diversity documented here means immunization must be embedded in a broader strategy that compliments vaccination with improved clinical care, antimicrobial stewardship and robust infection control.

These recommendations are likely to be relevant beyond Bangladesh and should inform intervention design across south Asia.

## Methods

### Ethics Statement

This study analyzes *KpSC* isolates, along with associated clinical and laboratory metadata, collected as part of the PneumoADIP study (2004-2008) and the Invasive Bacterial Disease (IBD) study (2009-2021). Ethical approvals for data collection, analysis and pathogen sequencing were obtained from the Ethical Review Board of the Bangladesh Institute of Child Health (now known as Bangladesh Shishu Hospital & Institute). All procedures were conducted in accordance with the National Ethical Guidelines for Health Research in Bangladesh issued by the Bangladesh Medical Research Council. We obtained written informed consent from the guardians of children admitted to the study hospitals, from whom blood and/or cerebrospinal fluid (CSF) samples were collected.

### Description of the study site

From 2004-2021, blood and/or CSF specimens were collected at four hospitals in Bangladesh: a) Bangladesh Shishu Hospital and Institute (BSHI, formerly known as Dhaka Shishu Hospital), a 650-bed tertiary level hospital based in Dhaka; b) Dr. M R Khan Shishu Hospital and Institute of Child Health (MRKSH), a 250-bed primary - secondary level hospital in Dhaka; c) Chattogram Maa-O Shishu Hospital (CMOSH), a 835-bed primary - tertiary level hospital in Chattogram, and d) Kumudini Women’s Medical College and Hospital (KWMCH), a 1050-bed based primary-tertiary level hospital based in rural town Mirzapur. BSHI and CMOSH operate Level III neonatal intensive care units, while MRKSH and KWMCH provide Level II neonatal care. Children hospitalized in these hospitals who matched the WHO-defined clinical criteria for pneumonia, severe pneumonia, severe disease, and meningitis were evaluated by trained research physicians. Blood and/or CSF was drawn for culture at the treating physicians’ discretion. Demographic (age and sex) and clinical (symptoms, hospital outcome, admission date, date of sample collection etc.) data were obtained for all enrolled patients (Sup Table 1). Children were enrolled upon informed written consent if they met the inclusion criteria.

### Laboratory procedures

Blood and/or CSF samples from patients were incubated in tryptic-soy broth and incubated at 37°C for overnight then sub-cultured on blood agar, chocolate agar and MacConkey agar media from 2004-2015. We introduced BACTEC for blood cultures in BSHI, KWMCH and MRKSH, in 2016 and in CMOSH the following year; since then, blood specimens were incubated in BACTEC culture systems for up to 120 hours, and positive samples were subsequently sub-cultured on blood agar, chocolate agar and MacConkey agar media. Upon growth, biochemical tests including, indole, urease, and citrate utilization tests were performed to identify the genus and species. Isolates with mucoid colony morphology, lactose fermentation on MacConkey agar, and a biochemical profile of oxidase-negative, indole-negative, urease-positive, and citrate-positive were confirmed as KpSC and stored at -80°C in STGG media (skim milk, tryptone, glucose, and glycerin). These stocks served as the source for all subsequent experiments performed in the study.

Antimicrobial susceptibility testing was performed using antibiotic discs by Kirby-Bauer testing for: meropenem, imipenem, ceftriaxone, ampicillin, gentamicin, netilmicin, ciprofloxacin, amikacin, chloramphenicol, and cotrimoxazole. Tests were interpreted according the Clinical and Laboratory Standards Institute (CLSI) 2025 antibiotic susceptibility testing clinical breakpoints^19^.

From 2011 to 2019, the microbiology laboratories, as part of the WHO’s Invasive Bacterial Vaccine-Preventable Diseases (IB-VPD) surveillance, underwent external quality assurance from UK-NEQAS.

### Sample selection criteria for whole genome sequencing (WGS)

Approximately a third of KpSC isolates from children under the age of 2 months, were randomly selected, keeping the distribution of isolates selected per year consistent. For each calendar year, all eligible isolates were enumerated, and a target proportion (∼33%) was selected using computer-generated random numbers, ensuring that the temporal distribution of sequenced isolates closely reflected the annual distribution of all culture-confirmed cases. These selected isolates were re-cultured from biobank, biochemically re-confirmed and sub-cultured on MacConkey agar media prior to DNA extraction and whole genome sequencing.

To contextualize the selected isolates with other KpSC isolates from Bangladesh, in addition to the isolates recovered in this study, we randomly selected 100 of 160 (62%) KpSC isolates for WGS that were stored in our biobank collected between 2004-2008 from three additional hospitals that followed the same inclusion criteria: a) Dhaka Medical College and Hospital, Dhaka, b) Chattogram Medical College and Hospital, Chattogram, and c) Sir Salimullah Medical College and Hospital, Dhaka as part of the pneumoADIP study.

### Genomic DNA extraction and sequencing

Total DNA from KpSC colonies was extracted using the QIAGEN DNA Mini extraction kit (Cat: 56304, Qiagen, Hilden, Germany). Sequencing libraries were prepared from extracted DNA using NEBNext Ultra II FS DNA Library kit (Cat: E7805, New England Biolabs, MA, USA). Libraries were sequenced on an Illumina NEXTSeq2000 at the Child Health Research Foundation, Dhaka, Bangladesh (n = 528; 150bp PE) and on the HiSeq4000 platform at the University of Oxford, Oxford, UK (n = 71; 150bp PE).

### Genome assembly

Sequence data quality was assessed using FastQC v0.11.5^20^ and MultiQC v3^21^. Adapter trimming was performed with Trimmomatic v0.39^22^. *De novo* genome assembly was conducted using Unicycler v0.4.9^23^, evaluated using Quast v.5.2.0^24^, and annotated with Prokka v.1.14.6^25^. Kleborate v2.2^26^ was used to identify the sequence types (STs), AMR genes, K and O antigen types. All generated sequence data have been submitted to ENA (BioProject: PRJEB90555).

### Phylogenetic Analysis

For reference-based genome mapping, Bowtie2 v2.3.4.1 was employed to align all genomic data against the NCBI reference genome, *Klebsiella pneumoniae* HS11286 (Genbank accession: NC_016845.1)^27^. SAMtools v1.7.2 and BCFtools v1.7.2 were utilized for variant calling, filtering candidate SNPs for a minimum base quality of 20, a minimum of 5 high-quality reads, and greater than 75% read support (i.e., over 75% of the reads supporting the SNP relative to the reference)^28^. SNPs were further filtered to remove potential biases, including strand, mapping, and end biases (p-value < 0.001). Additionally, SNPs located within phage regions (identified by PHASTER^29^), repeat sequences (as noted in the reference GenBank), and recombination regions (detected by Gubbins v2.3.1) were excluded^30^. The resulting filtered SNPs were then converted into a SNP alignment for maximum likelihood phylogenetic tree (MLT) construction using RAxML v8.2.11 (model: GTRGAMMA; 100 bootstraps)^31^. The alignment had 82,669 SNP positions from 472 *Klebsiella pneumoniae* isolates in our study. Tree visualization and annotation was done using iTOL v6^32^.

For global comparison of the STs of the *K. pneumoniae* isolates (n = 472) sequenced in this study, publicly available *K. pneumoniae* genomes (n = 32,656) in PathogenWatch^33^ (retrieved on November 29, 2023) were used.

### Statistical analysis

Statistical analyses were performed using custom scripts in R (version 4.3.1). The odds ratio was calculated with using the *oddsratio* package, while the figures were generated using the *ggplot2* and *Rcolorbrewer* and *ggsci* packages. Global and Bangladesh-specific maps were made using the packages *rnaturalearth, ggspatial, maps* and *scatterpie.* For ST11 geographic analysis, upazila, village or union information was used instead of the exact address obtained during data collection. Hospital outcome was classified as died, discharged, or left against medical advice (LAMA).

Hospital case-fatality rate (CFR) was defined as the proportion of culture-confirmed KpSC cases that died during hospitalization, thus LAMA and discharged cases were included in the primary CFR denominator. To evaluate the potential impact of LAMA on mortality estimates, we performed a bounded sensitivity analysis under three assumptions: (i) a primary analysis excluding LAMA cases (death / [death + discharged]); (ii) a lower-bound estimate, assuming all LAMA cases survived (death / total cases); and (iii) an upper-bound estimate, assuming all LAMA cases died ([death + LAMA] / total cases). Year-specific mortality estimates, and 95% confidence intervals were calculated for each scenario.

To assess the association between carbapenem resistance and in-hospital mortality, we fitted logistic regression models with death during hospitalization as the binary outcome. Carbapenem resistance was the primary exposure of interest. Multivariable models were adjusted for available demographic, temporal, and hospital-level covariates, including age group (neonate [≤28 days] vs older), sex, hospital site, timing of culture collection relative to admission, and admission year or epoch (2004-2015 vs 2016-2021) to account for secular trends. To evaluate robustness, we fitted a series of models adjusting for individual covariates and all possible combinations thereof. Adjusted odds ratios and 95% confidence intervals were reported.

## Data availability

All data supporting the findings of this study are available within the paper and its Supplementary Information. All whole genome sequencing data are available on ENA (BioProject: PRJEB90555).

## Acknowledgements

We thank all study participants and their families for their willingness to contribute to this research. We are grateful to the physicians, nurses, data collectors, and laboratory personnel at the participating hospitals for their dedicated patient care and for generating the clinical and laboratory data used in this study. We thank the funder, Gates Foundation (INV-023821 to Dr. Senjuti Saha) of supporting this study. The funder had no role in the study design, data collection, analysis, interpretation, or the decision to publish the findings.

## Author Contributions

SS and SKS contributed to conceptualization, funding acquisition, study design, implementation, and monitoring. SS, YH, AMT and NK contributed to data analysis, preparation of figures, drafting the initial manuscript, and manuscript editing. MSI contributed to data analysis and coordination of the study. HR and MdH coordinated the blood culture, antimicrobial susceptibility, and other laboratory testing. AAT, SG and TJ performed DNA extraction and whole genome sequencing. YH, AMT and MdMH conducted the comparative genomic analysis. ZBA, BH, ShS, MJU and ABJ coordinated and trained doctors and nurses, and assisted in the implementation of the study. RCD performed data cleaning. MS, NKG, AKMS, NS, SKB, FA, WA, MMH, MH and ASMNU contributed to study management and patient data analysis. All authors reviewed the manuscript and agreed on the contents. The corresponding authors had full access to all the data in the study and had final responsibility for the decision to submit for publication. YH, NK, and AMT directly accessed and verified the data. All authors had access to de-identified data.

