## Supplementary Information for "Escalating burden and mortality of carbapenem-resistant *Klebsiella pneumoniae* species complex infections in Bangladeshi infants"

*Hooda et al*

### Supplementary text descriptions of methods

#### Text S1. Inclusion criterion used in the study.

Patients admitted to the pediatric wards selected for this study were identified using the ward admission logbook. Study physicians conducted a detailed clinical examination and took a thorough medical history. Children aged 0–59 months admitted to the selected wards were evaluated using the RSV hospital-based surveillance case definition. Children hospitalized with a respiratory infection, defined as having cough or shortness of breath, with an onset within the last 10 days, were eligible to be enrolled in the study. In addition, infants younger than six months were also eligible if they presented with apnoea, which is characterized by a temporary cessation of breathing from any cause and/or sepsis (defined as fever (temperature of 37.5 °C or above) or hypothermia (temperature less than 35.5 °C), with indications of shock (lethargy, fast breathing, cold skin, prolonged capillary refill, or a fast, weak pulse). Once a child met the criteria, caregivers were counseled about the study, including its objectives, procedures, and potential risks and benefits. Written consent was mandatory for participation, covering both data collection and nasopharyngeal swab sampling. Following consent, the patient was enrolled, and data was recorded using a standardized, tablet-based system. Clinical follow-up was conducted daily for each enrolled patient until discharge, referral, death, or the family left against medical advice. Physicians and research assistants documented clinical signs, laboratory findings, treatment details, and final outcomes.

### Supplementary Figures

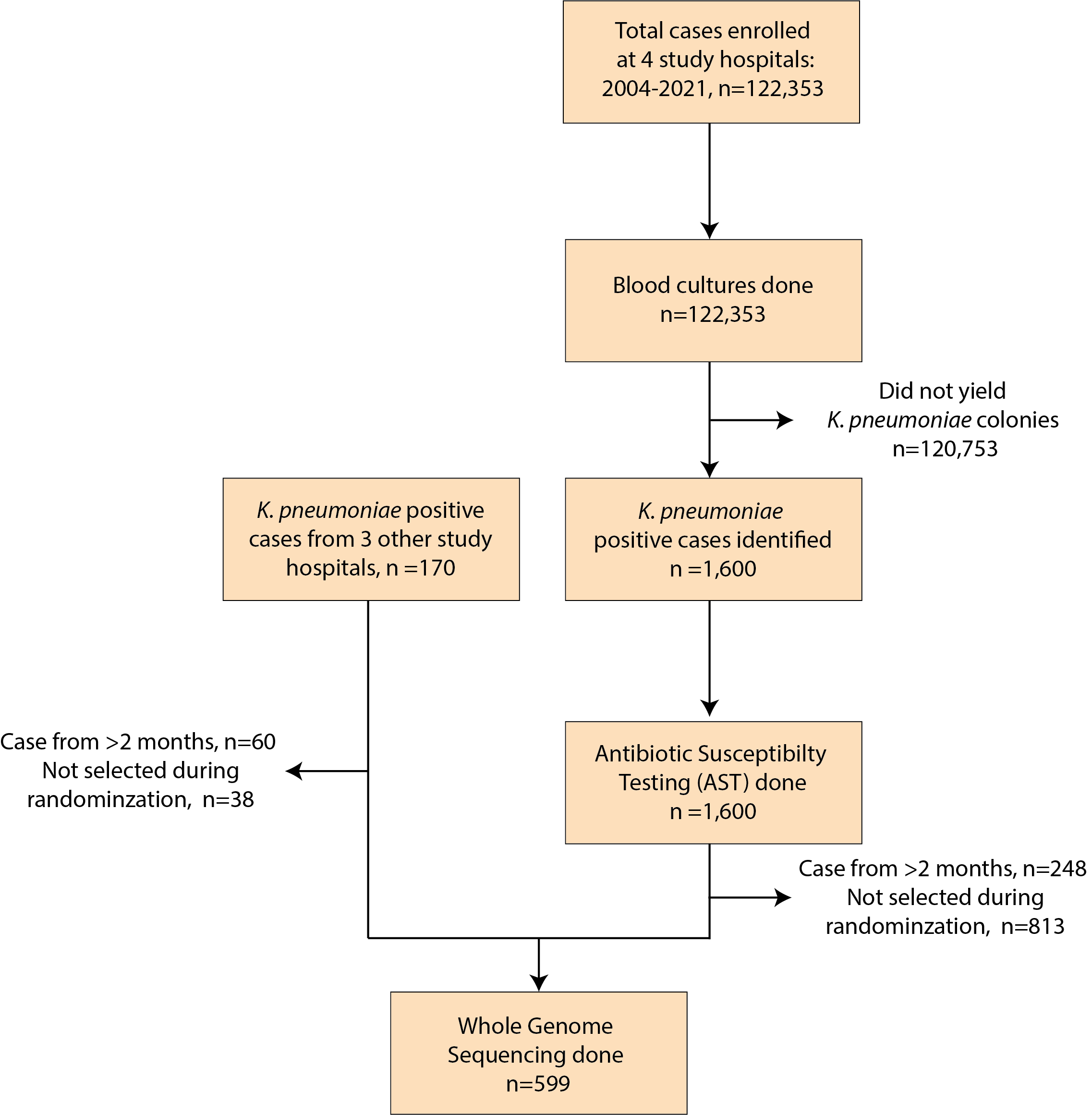

#### Supplementary Figure 1: Flow chart of the study

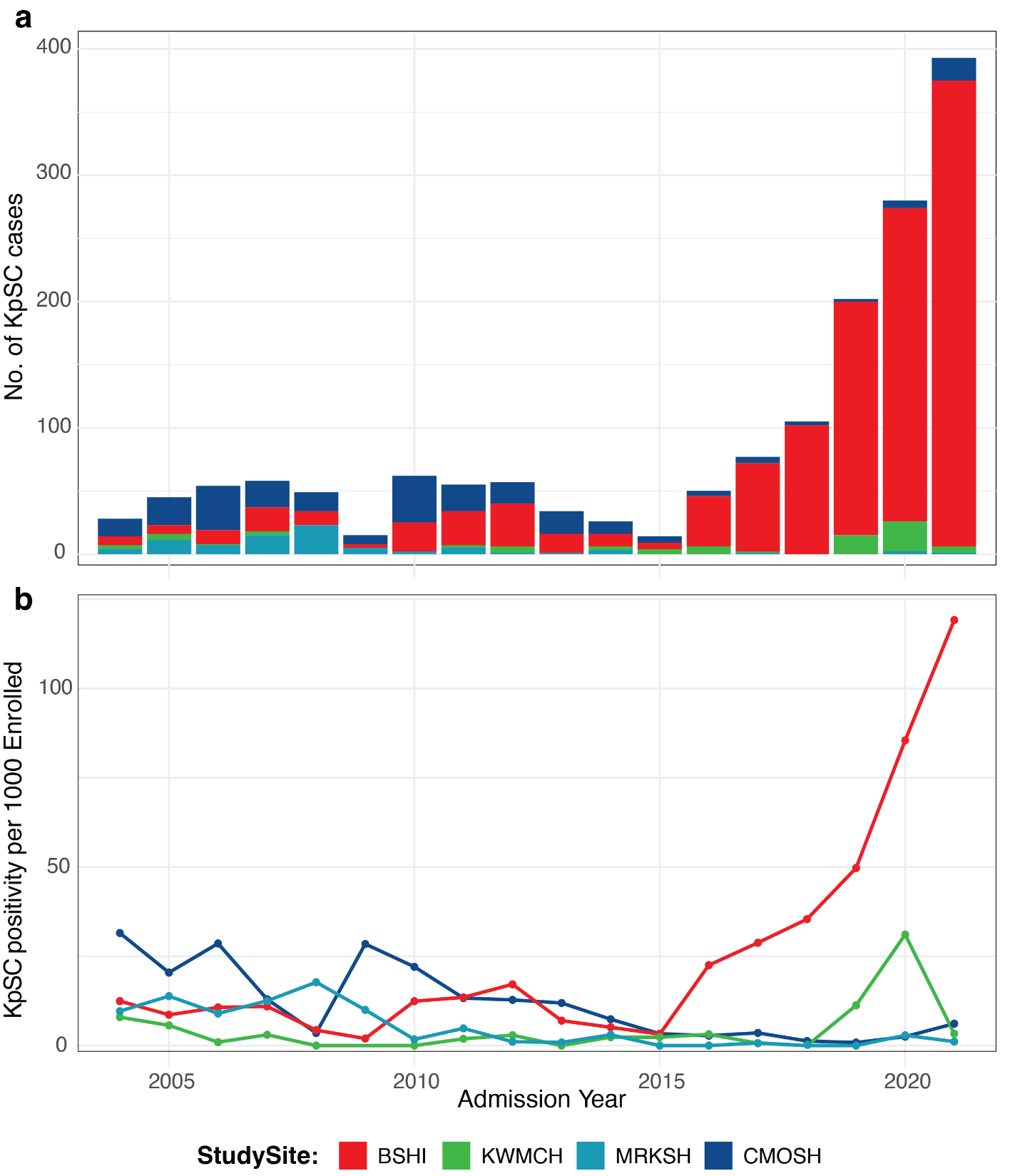

Supplementary Figure 2: Number and positivity rate of KpSC isolates at the study hospitals, 2004-2021. a, Number of KpSC isolates sub-divided by the study site of isolation. b, Trends in KpSC isolation rates at the study sites.

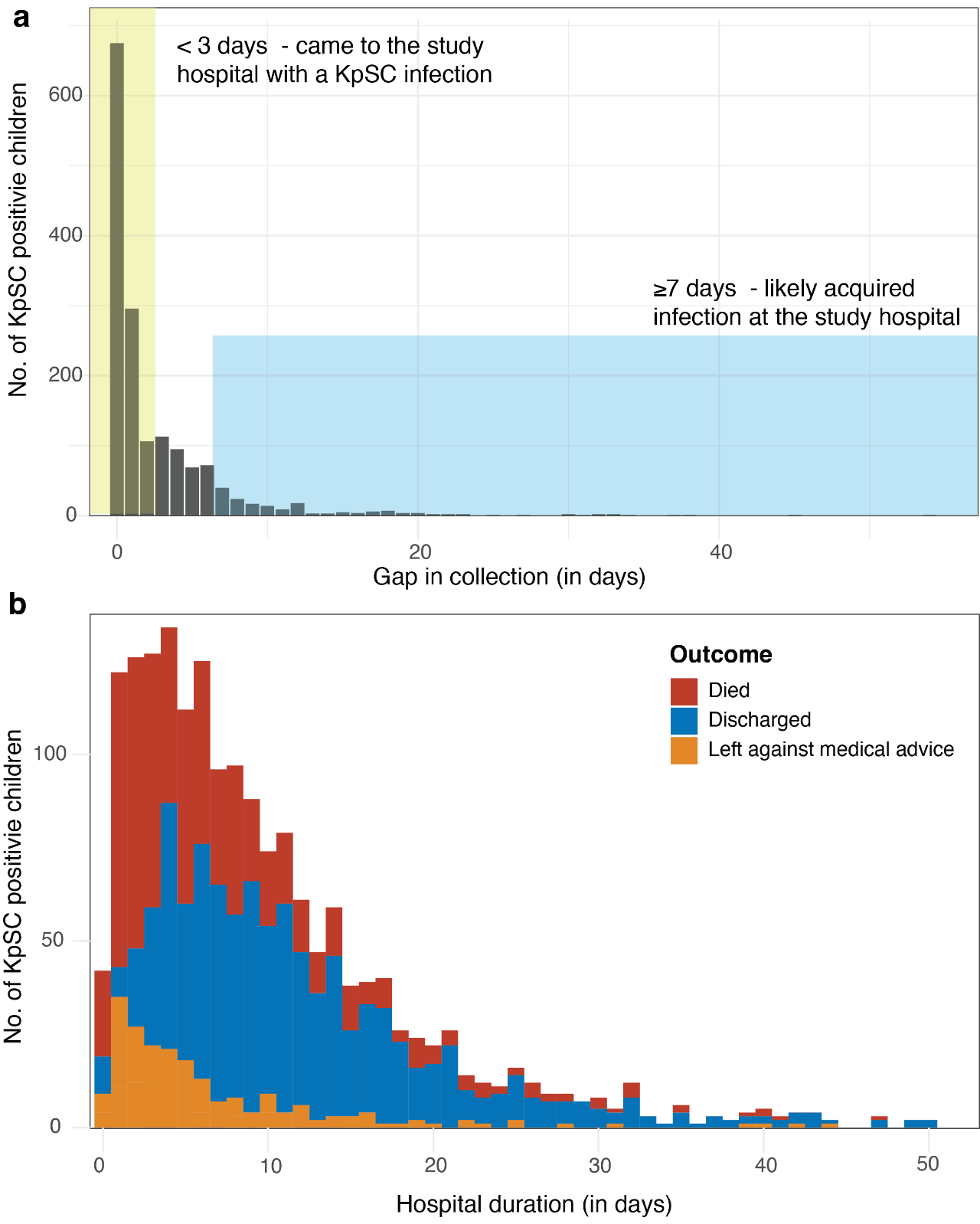

Supplementary Figure 3: Difference (in days) between hospital admission & blood draw and hospital stay for KpSC positive cases. **a,** Gap in collection is defined as the difference (in days) between hospital admission and blood draw. 67% of Kpsc positive infections were identified through blood cultures less than 72 hours (0-2 days) from admission; these were likely acquired outside the study hospitals. **b,** Hospital duration of children with KpSC infections that presented at the study hospital. 64% of children died within 5 days of admission.

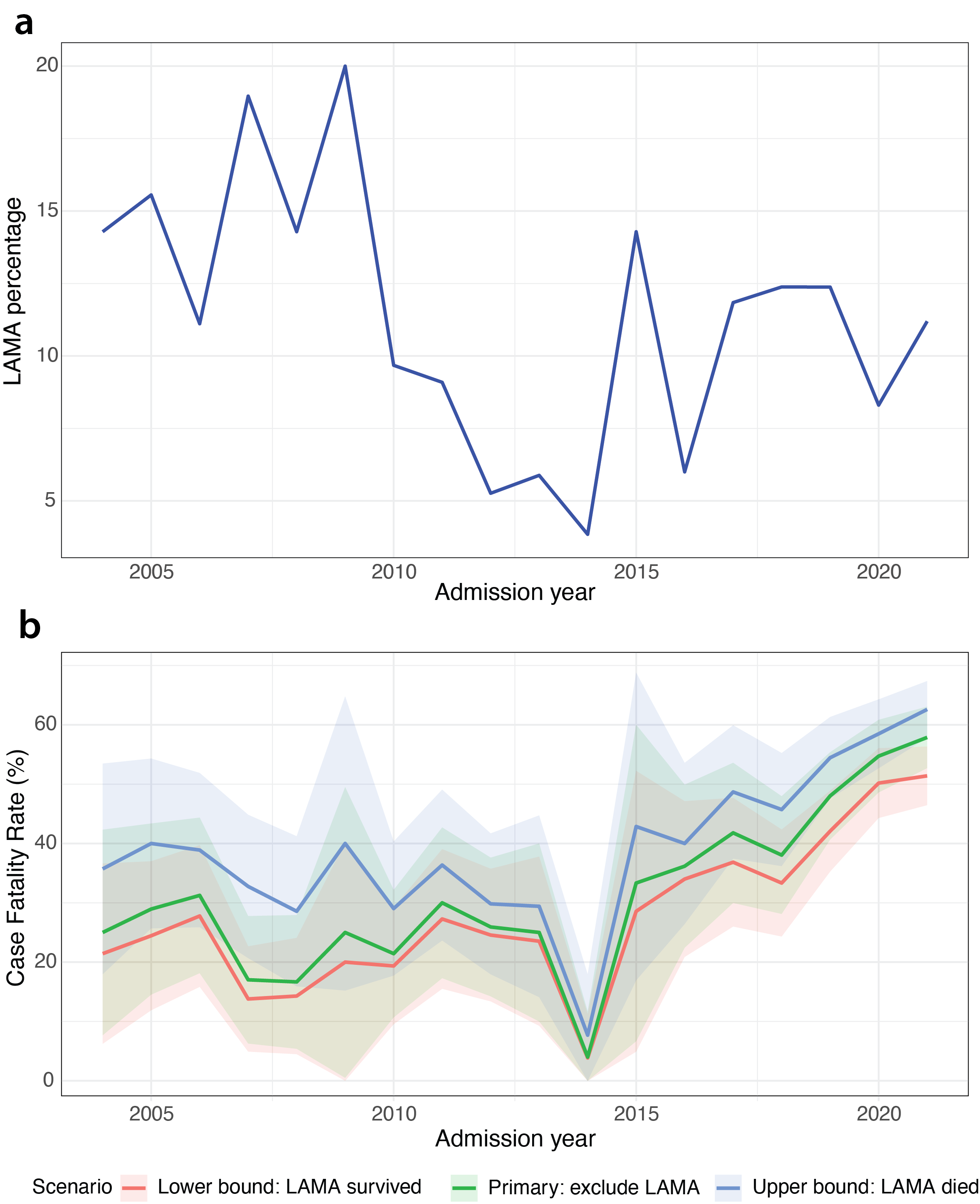

Supplementary Figure 4: Lost to follow-up (LAMA) rates. **a,** Percentage of annual cases where the hospital outcome was LAMA. **b,** Sensitivity analysis to investigate the impact of LAMA on trends in the study under three assumptions: (i) a primary analysis excluding LAMA cases (death / [death + discharged]); (ii) a lower-bound estimate, assuming all LAMA cases survived (death / total cases); and (iii) an upper-bound estimate, assuming all LAMA cases died ([death + LAMA] / total cases). Year-specific mortality estimates, and 95% confidence intervals are shown for each scenario.

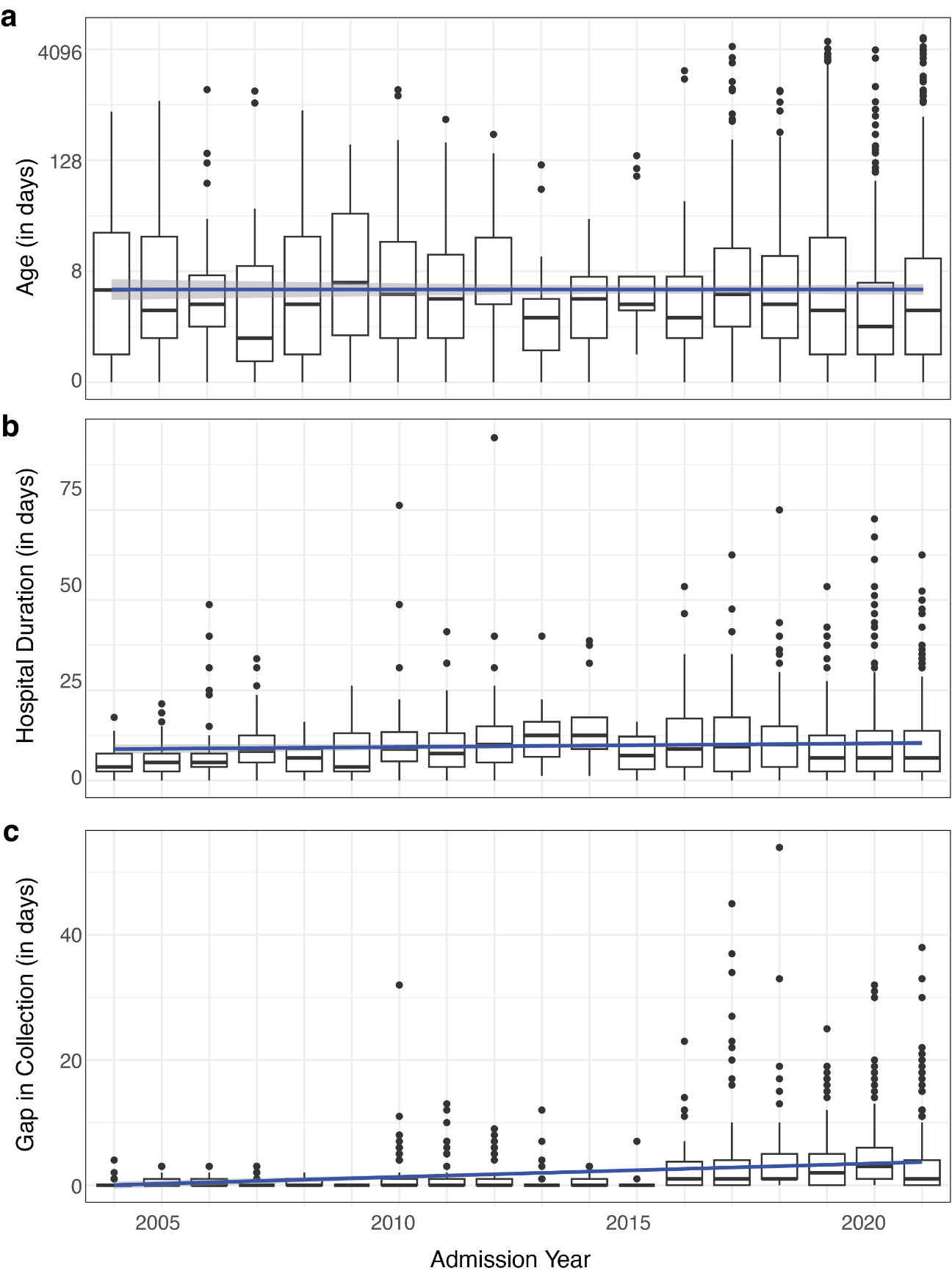

Supplementary Figure 5: Trends in a, age (in days), b, hospital stay after blood draw (Hospital Duration, in days) and c, Gap in collection (in days) amongst KpSC positive cases from 2004-2021. Blue line shows the linear trendline.

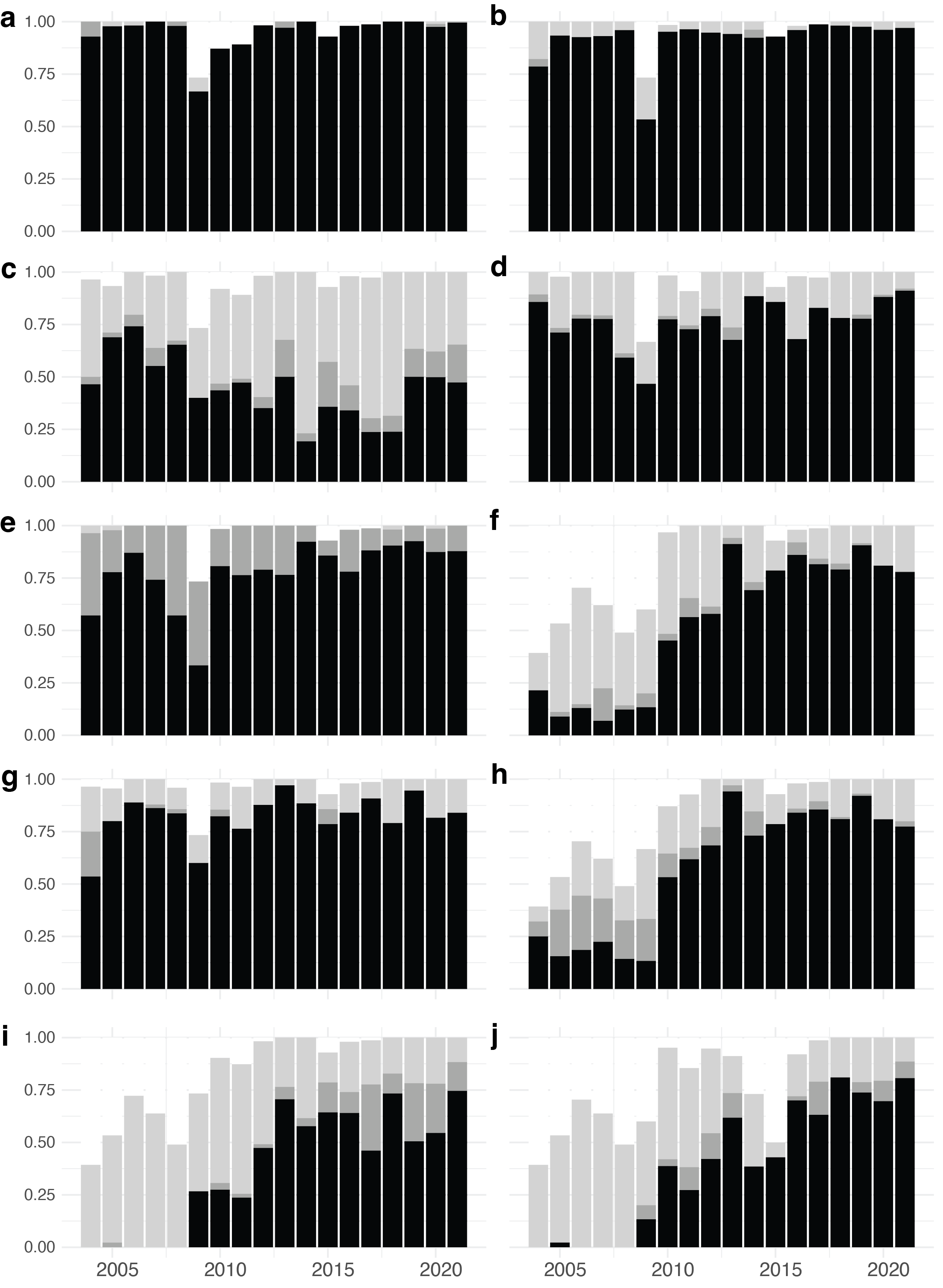

Supplementary Figure 6: Trends in antimicrobial susceptibility from 2004-2021 for 9 different antibiotics recommended by CLSI. Trends in resistance for **a,** Ampicillin, **b,** Ceftriaxone, **c,** Chloramphenicol, **d,** Cotrimoxazole, **e,** Ciprofloxacin, **f,** Amikacin, **g,** Gentamicin, **h,** Netilmicin, **i,** Imipenem, **j,** Meropenem

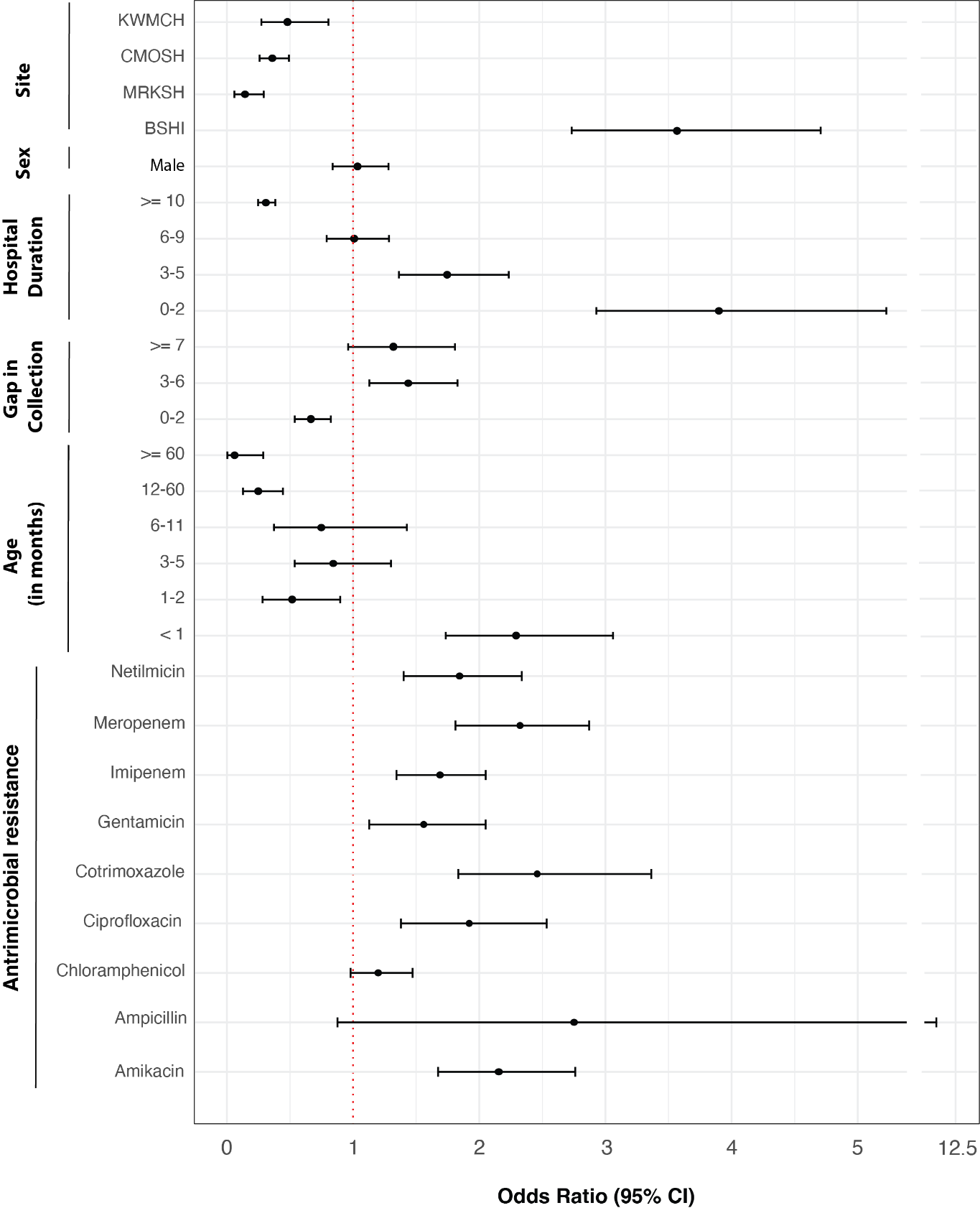

#### Supplementary Figure 7: Odds ratio (95% CI) of mortality stratified by site, sex, hospital duration (in days), gap collection (in days), age at admission (in months) and resistance against 10 different antibiotics recommended by CLSI.

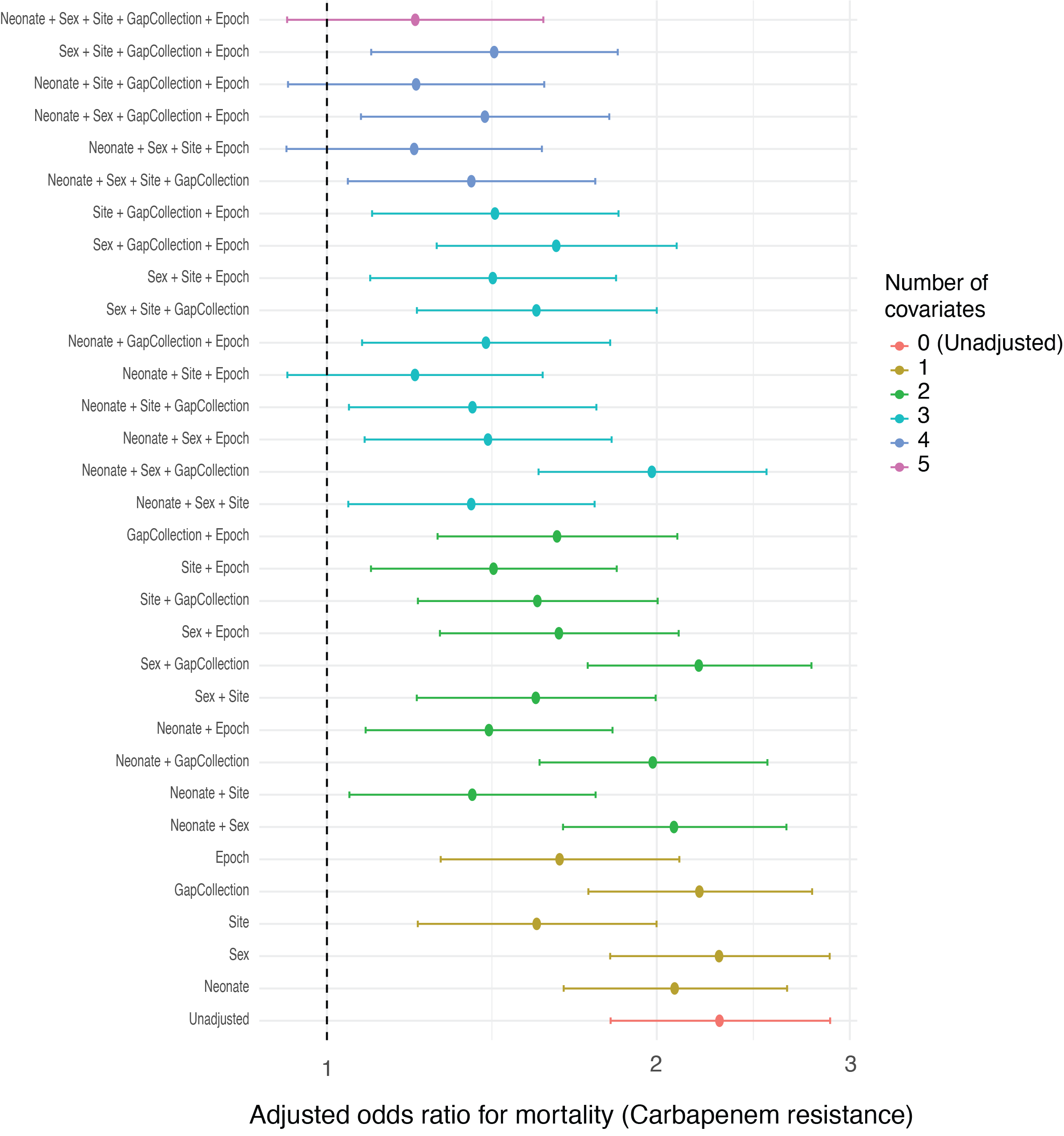

#### Supplementary Figure 8: Sensitivity analysis of the association between carbapenem resistance and in-hospital mortality under varying covariate adjustment sets.

Forest plot showing odds ratios (points) and 95% confidence intervals (horizontal bars) for mortality associated with carbapenem-resistant KpSC across progressively adjusted logistic regression models. Each row represents a distinct model including different combinations of covariates (neonatal status, sex, hospital site, gap in specimen collection, and admission epoch (2004-2015 vs 2016-2021)). Colors indicate the number of covariates included in each model, ranging from unadjusted (0 covariates) to fully adjusted (5 covariates). The vertical dashed line denotes the null value (odds ratio = 1).

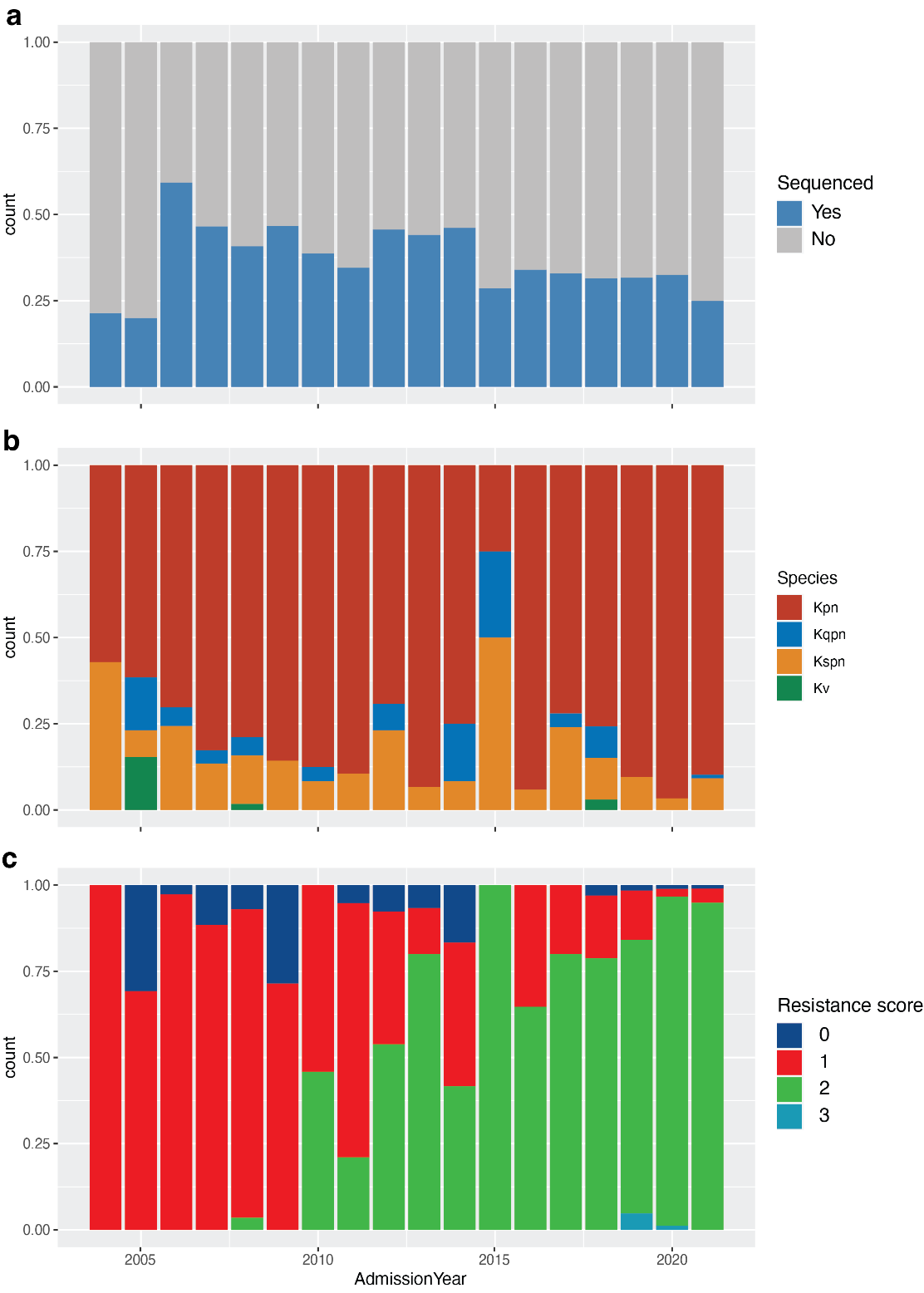

Supplementary Figure 9: Selection of strains for sequencing and trends in species and antibiotic resistance amongst KpSC isolates from 2004-2021. **a,** Strains were selected randomly from different years. **b,** Yearly trends in KpSC species identified. Kpn: *K. pneumoniae*, Kqpn: *K. quasipneumoniae subsp quasipneumoniae*, Kspn: *K. quasipneumoniae subsp similipneumoniae*, Kv: *K. variicola*. **c,** Yearly trends in isolation of genomic resistance predicted by *Kleborate*, where 0 – no resistance to extended spectrum beta-lactamases (ESBL), 1 – resistance to ESBLs, 2 – resistance to ESBLs + Carbapenems, 3 – resistance to ESBLs + Carbapenems + Colistin.

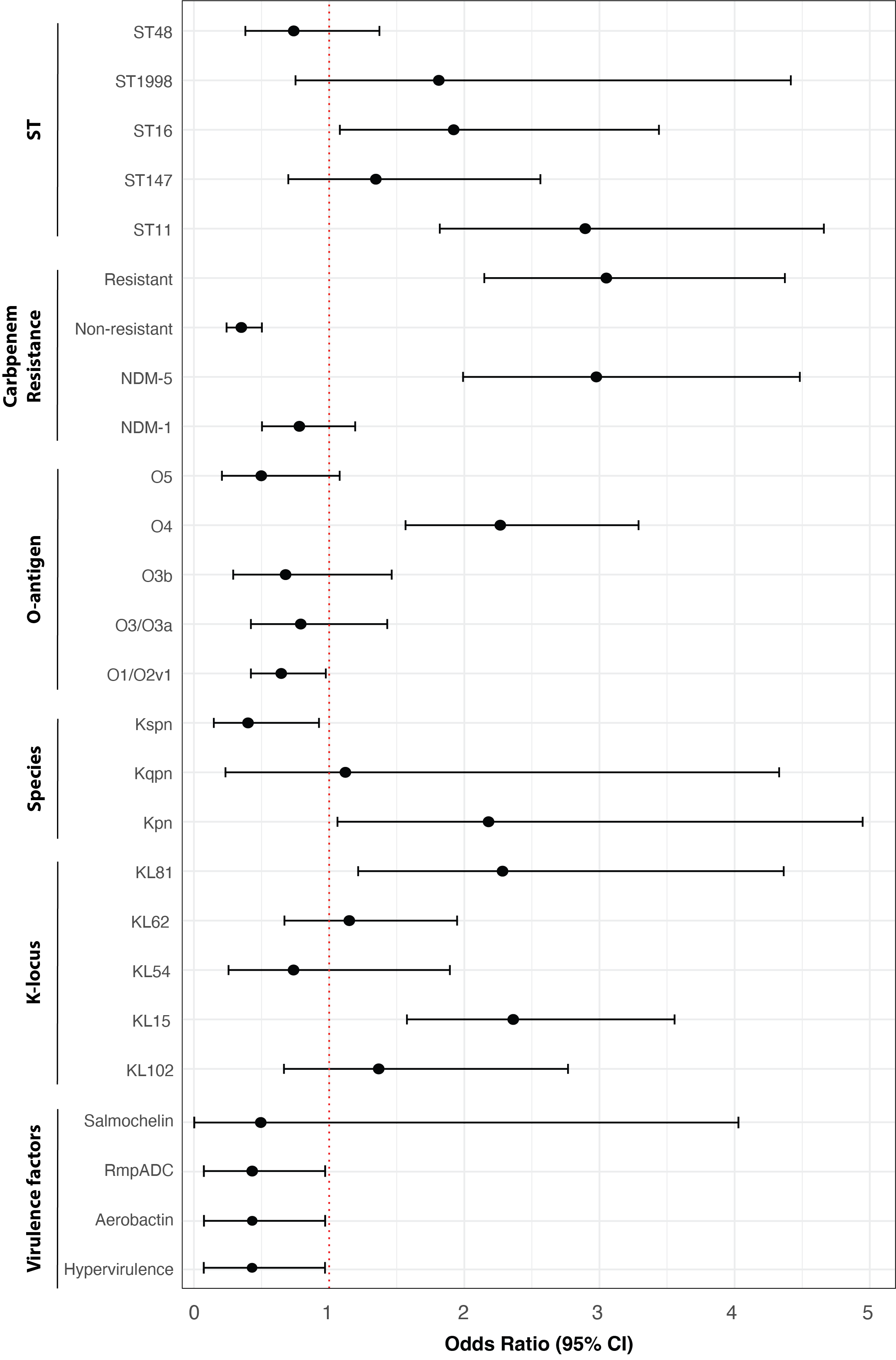

Supplementary Figure 10: Odds ratio of mortality for different KpSC species, carbapenem resistance markers and the five most common STs, O-types, K-types and virulence factors. Odds ratio of carbapenem resistance, non-resistance and resistance due to *ndm-5* and *ndm-1* gene is shown. Kpn: *K. pneumoniae*, Kqpn: *K. quasipneumoniae subsp quasipneumoniae*, Kspn: *K. quasipneumoniae subsp similipneumoniae*, Kv: *K. variicola*. Hypervirulence is defined as based on the Kleborate virulence score >= 3.

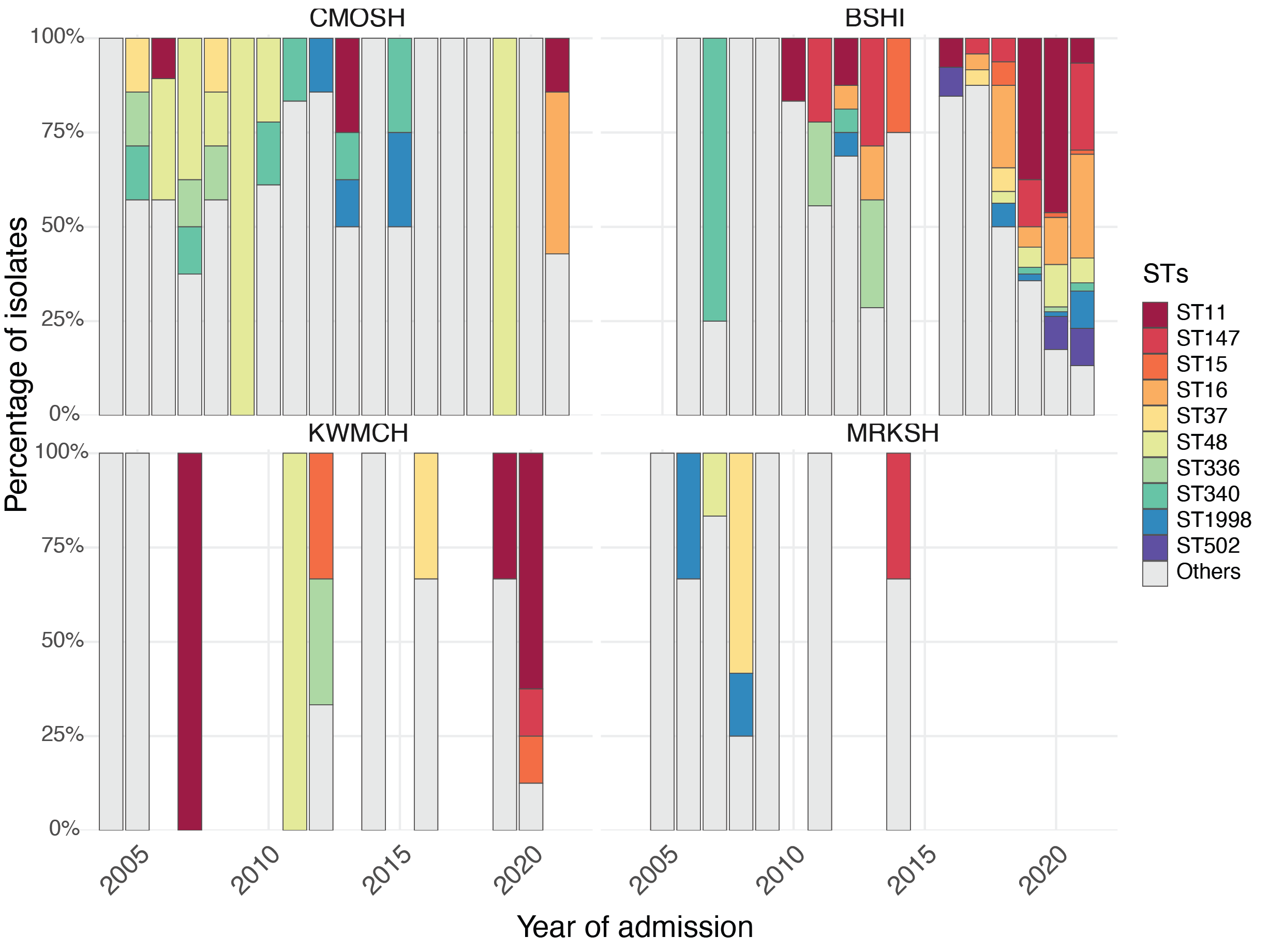

#### Supplementary Figure 11: Sequence-type distribution across the four hospital sites

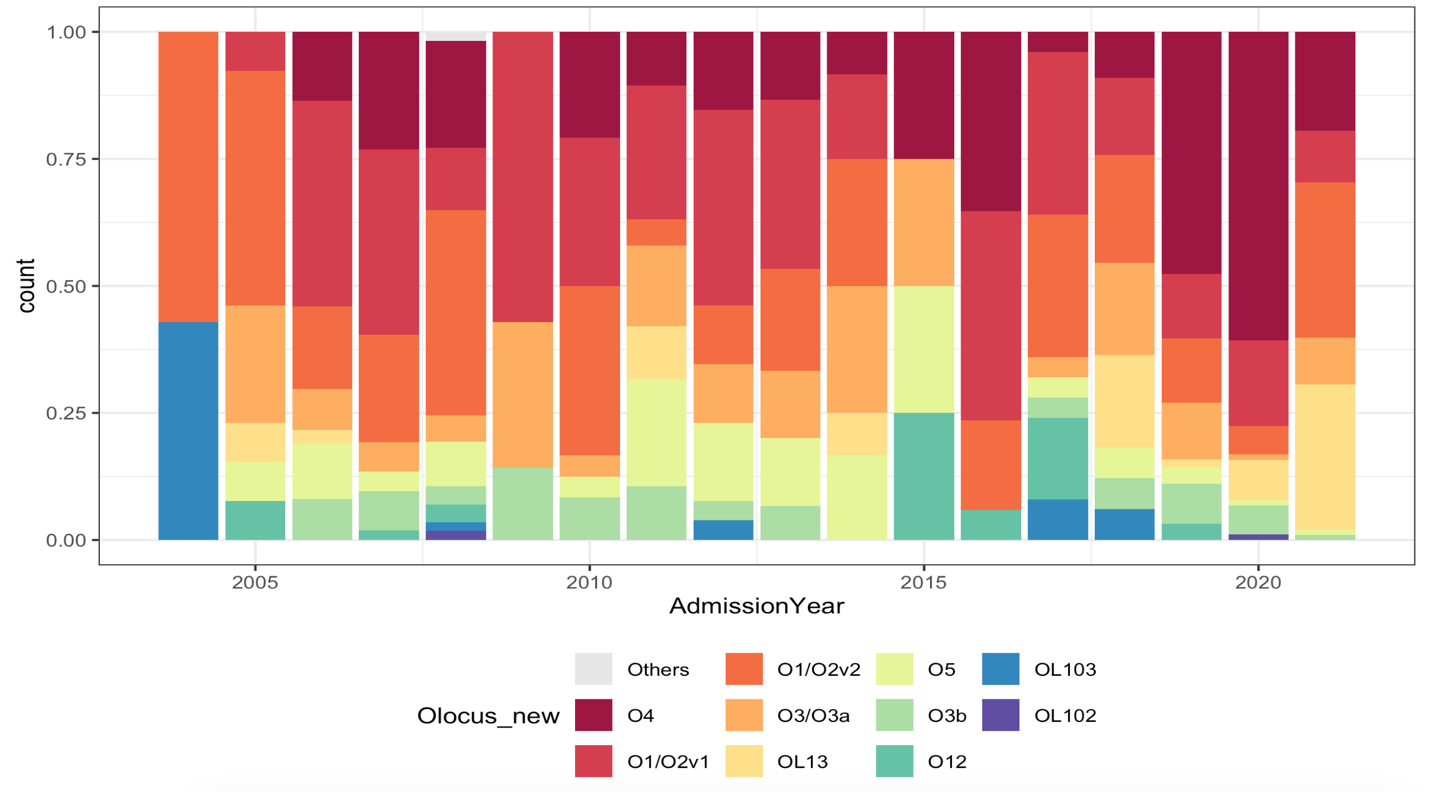

#### Supplementary Figure 12: Trends in ten most frequent O-types from 2004-2021

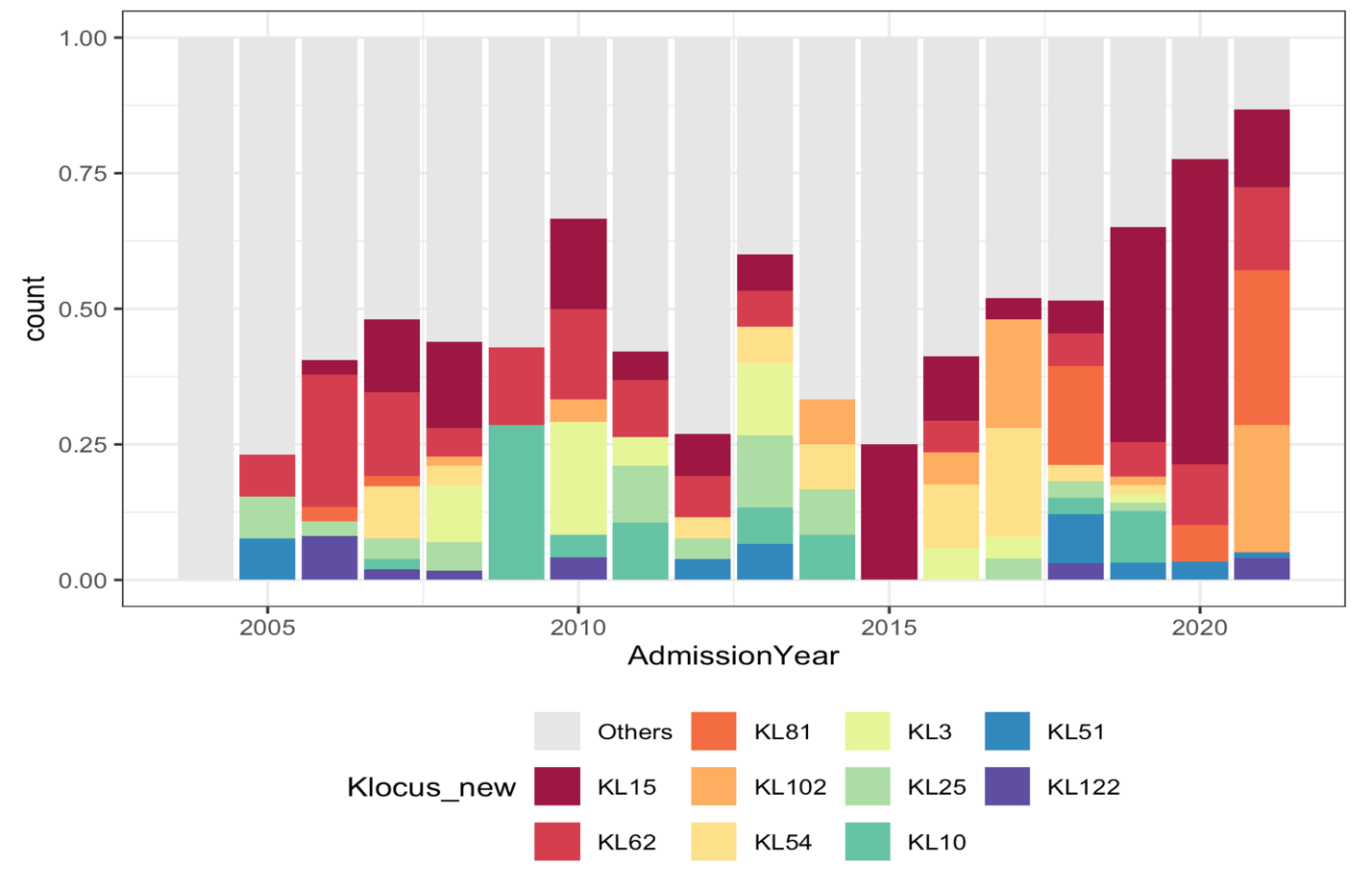

#### Supplementary Figure 13: Trends in ten most frequent K-locus from 2004-2021

### Supplementary Tables

| **All Analyzed- Klebsiella (1600)** | | |
| --- | --- | --- |
| **Clinical Features** | **Frequency** | **Percentage (%)** |
| **Sex** |  |  |
| Male | 1,044 | 65.25 |
| Female | 556 | 34.75 |
| **StudySite** |  |  |
| Dhaka Shisu Hospital (DSH) | 1182 | 73.88 |
| Chittagong Ma O Shisu Hospital (CMOSH) | 260 | 16.25 |
| Shisu Shastho Foundation (SSF) | 81 | 5.06 |
| Kumudini Womens Medical College Hospital (KWMCH) | 77 | 4.81 |
| **Admission year** |  |  |
| 2004 | 28 | 1.75 |
| 2005 | 45 | 2.81 |
| 2006 | 54 | 3.38 |
| 2007 | 58 | 3.63 |
| 2008 | 49 | 3.06 |
| 2009 | 15 | 0.94 |
| 2010 | 62 | 3.88 |
| 2011 | 55 | 3.44 |
| 2012 | 57 | 3.56 |
| 2013 | 34 | 2.13 |
| 2014 | 26 | 1.63 |
| 2015 | 14 | 0.88 |
| 2016 | 50 | 3.13 |
| 2017 | 76 | 4.75 |
| 2018 | 105 | 6.57 |
| 2019 | 202 | 12.63 |
| 2020 | 277 | 17.31 |
| 2021 | 393 | 24.56 |
| **Age in Month (at Admission Date)** |  |  |
| 0m | 1297 | 81.06 |
| 1m | 55 | 3.44 |
| 2-5m | 92 | 5.75 |
| 6-11m | 43 | 2.69 |
| 12-23m | 39 | 2.44 |
| 24-35m | 17 | 1.06 |
| 36-47m | 16 | 1 |
| 48-59m | 11 | 0.69 |
| >59m | 30 | 1.88 |
| **Age in Month (at Specimen Collection Date)** |  |  |
| 0m | 1291 | 80.69 |
| 1m | 61 | 3.81 |
| 2-5m | 92 | 5.75 |
| 6-11m | 43 | 2.69 |
| 12-23m | 38 | 2.38 |
| 24-35m | 17 | 1.06 |
| 36-47m | 16 | 1 |
| 48-59m | 12 | 0.75 |
| >59m | 30 | 1.88 |
| **Case Definition** |  |  |
| Pneumonia | 28 | 1.75 |
| Severe Pneumonia | 471 | 29.44 |
| Meningitis | 99 | 6.19 |
| Very Severe Disease | 573 | 35.81 |
| Enteric Fever | 10 | 0.63 |
| Not Eligible | 393 | 24.56 |
| Missing | 26 | 1.63 |
| **Specimen Type** |  |  |
| Blood | 1594 | 99.63 |
| CSF | 5 | 0.31 |
| Both (Blood & CSF) | 1 | 0.06 |
| **Hospital Outcome** |  |  |
| Discharged | 816 | 51 |
| Died | 610 | 38.13 |
| Left against medical advice | 174 | 10.87 |
| **Hospital Duration** |  |  |
| 0-2d | 241 | 15.07 |
| 3-5d | 323 | 20.2 |
| 6-10d | 425 | 26.58 |
| 11-15d | 272 | 17.01 |
| 16-28d | 256 | 16.01 |
| >28d | 82 | 5.13 |
| **Difference between Admission Date and Specimen Collection Date** | |  |
| 0-2d | 1076 | 67.25 |
| >2d | 524 | 32.75 |
| **Final Diagnosis** |  |  |
| Systemic infections | 935 | 58.44 |
| Perinatal asphyxia | 512 | 32.00 |
| Preterm low-birth weight | 344 | 21.50 |
| Respiratory manifestation | 292 | 18.25 |
| Congenital Cardiovascular manifestation | 270 | 16.88 |
| Neonatal jaundice | 251 | 15.69 |
| Neurological manifestation | 49 | 3.06 |
| Febrile illness | 15 | 0.94 |
| Gastrointestinal manifestation | 12 | 0.75 |
| Genitourinary/Renal manifestation | 14 | 0.88 |
| Others | 1269 | 79.31 |

#### Supplementary Table 1: Summary of clinical and demographic data on the 1,600 KpSC positive cases analyzed in this study.

| **All Sequenced- Klebsiella (599)** | | |
| --- | --- | --- |
| **Clinical Features** | **Frequency** | **Percentage (%)** |
| **StudySite** |  |  |
| Dhaka Shisu Hospital (DSH) | 347 | 57.93 |
| Chittagong Ma O Shisu Hospital (CMOSH) | 120 | 20.03 |
| Shisu Shastho Foundation (SSF) | 32 | 5.34 |
| Kumudini Womens Medical College Hospital (KWMCH) | 28 | 4.67 |
| Chittagong Medical College Hospital (CMCH) | 38 | 6.34 |
| Dhaka Medical College Hospital (DMCH) | 23 | 3.84 |
| Sir Salimullah Medical Colllege Hospital (SSMCH) | 11 | 1.84 |
| **Admission year** |  |  |
| 2004 | 7 | 1.17 |
| 2005 | 13 | 2.17 |
| 2006 | 37 | 6.18 |
| 2007 | 52 | 8.68 |
| 2008 | 57 | 9.52 |
| 2009 | 7 | 1.17 |
| 2010 | 24 | 4.01 |
| 2011 | 19 | 3.17 |
| 2012 | 26 | 4.34 |
| 2013 | 15 | 2.5 |
| 2014 | 12 | 2 |
| 2015 | 4 | 0.67 |
| 2016 | 17 | 2.84 |
| 2017 | 25 | 4.17 |
| 2018 | 33 | 5.51 |
| 2019 | 64 | 10.68 |
| 2020 | 89 | 14.86 |
| 2021 | 98 | 16.36 |
| **Species** |  |  |
| Klebsiella pneumoniae | 503 | 83.97 |
| Klebsiella quasipneumoniae subsp. quasipneumoniae | 20 | 3.34 |
| Klebsiella quasipneumoniae subsp. Similipneumoniae | 72 | 12.02 |
| Klebsiella variicola subsp. variicola | 4 | 0.67 |
| **ST** |  |  |
| ST11 | 87 | 14.52 |
| ST16 | 51 | 8.51 |
| ST48 | 47 | 7.85 |
| ST147 | 40 | 6.68 |
| ST1998 | 21 | 3.51 |
| ST340 | 20 | 3.34 |
| ST502 | 17 | 2.84 |
| ST15 | 13 | 2.17 |
| ST37 | 13 | 2.17 |
| ST336 | 12 | 2 |
| ST490 | 11 | 1.84 |
| ST307 | 10 | 1.67 |
| ST14 | 9 | 1.5 |
| ST1473 | 9 | 1.5 |
| ST626 | 9 | 1.5 |
| ST152 | 8 | 1.34 |
| ST437 | 8 | 1.34 |
| ST70 | 8 | 1.34 |
| ST101 | 6 | 1 |
| ST394 | 6 | 1 |
| ST2482-1LV | 5 | 0.83 |
| ST530 | 5 | 0.83 |
| ST17 | 4 | 0.67 |
| ST1933 | 4 | 0.67 |
| ST231 | 4 | 0.67 |
| ST273 | 4 | 0.67 |
| ST39 | 4 | 0.67 |
| ST54 | 4 | 0.67 |
| ST107 | 3 | 0.5 |
| ST1991-1LV | 3 | 0.5 |
| ST23 | 3 | 0.5 |
| ST25 | 3 | 0.5 |
| ST2805 | 3 | 0.5 |
| ST29 | 3 | 0.5 |
| ST334 | 3 | 0.5 |
| ST5760 | 3 | 0.5 |
| ST705 | 3 | 0.5 |
| ST711 | 3 | 0.5 |
| ST841 | 3 | 0.5 |
| ST105 | 2 | 0.33 |
| ST1161 | 2 | 0.33 |
| ST1224 | 2 | 0.33 |
| ST1490 | 2 | 0.33 |
| ST1584-1LV | 2 | 0.33 |
| ST1681-1LV | 2 | 0.33 |
| ST1697 | 2 | 0.33 |
| ST1822 | 2 | 0.33 |
| ST2258 | 2 | 0.33 |
| ST2661 | 2 | 0.33 |
| ST3175-1LV | 2 | 0.33 |
| ST334-1LV | 2 | 0.33 |
| ST337 | 2 | 0.33 |
| ST34 | 2 | 0.33 |
| ST378 | 2 | 0.33 |
| ST420 | 2 | 0.33 |
| ST441-1LV | 2 | 0.33 |
| ST45 | 2 | 0.33 |
| ST540 | 2 | 0.33 |
| ST76 | 2 | 0.33 |
| ST873 | 2 | 0.33 |
| ST1019 | 1 | 0.17 |
| ST1040 | 1 | 0.17 |
| ST1040-1LV | 1 | 0.17 |
| ST1077-2LV | 1 | 0.17 |
| ST111-1LV | 1 | 0.17 |
| ST1304 | 1 | 0.17 |
| ST1309 | 1 | 0.17 |
| ST138 | 1 | 0.17 |
| ST1408 | 1 | 0.17 |
| ST1416 | 1 | 0.17 |
| ST1601-3LV | 1 | 0.17 |
| ST163 | 1 | 0.17 |
| ST1647-2LV | 1 | 0.17 |
| ST1655 | 1 | 0.17 |
| ST1676 | 1 | 0.17 |
| ST1681-3LV | 1 | 0.17 |
| ST1685 | 1 | 0.17 |
| ST1804 | 1 | 0.17 |
| ST185 | 1 | 0.17 |
| ST1855 | 1 | 0.17 |
| ST1887-2LV | 1 | 0.17 |
| ST197 | 1 | 0.17 |
| ST2054 | 1 | 0.17 |
| ST208 | 1 | 0.17 |
| ST2136 | 1 | 0.17 |
| ST22 | 1 | 0.17 |
| ST22-1LV | 1 | 0.17 |
| ST2218-1LV | 1 | 0.17 |
| ST2301 | 1 | 0.17 |
| ST234 | 1 | 0.17 |
| ST234-1LV | 1 | 0.17 |
| ST2389 | 1 | 0.17 |
| ST2478 | 1 | 0.17 |
| ST252 | 1 | 0.17 |
| ST2558 | 1 | 0.17 |
| ST2674 | 1 | 0.17 |
| ST268 | 1 | 0.17 |
| ST2727-1LV | 1 | 0.17 |
| ST2941 | 1 | 0.17 |
| ST297 | 1 | 0.17 |
| ST3266 | 1 | 0.17 |
| ST3375-1LV | 1 | 0.17 |
| ST35 | 1 | 0.17 |
| ST3512-1LV | 1 | 0.17 |
| ST3623 | 1 | 0.17 |
| ST3701 | 1 | 0.17 |
| ST38 | 1 | 0.17 |
| ST3871 | 1 | 0.17 |
| ST392 | 1 | 0.17 |
| ST395 | 1 | 0.17 |
| ST399-1LV | 1 | 0.17 |
| ST4085 | 1 | 0.17 |
| ST410 | 1 | 0.17 |
| ST411 | 1 | 0.17 |
| ST414 | 1 | 0.17 |
| ST429 | 1 | 0.17 |
| ST43 | 1 | 0.17 |
| ST4406 | 1 | 0.17 |
| ST4511 | 1 | 0.17 |
| ST476 | 1 | 0.17 |
| ST477 | 1 | 0.17 |
| ST480-2LV | 1 | 0.17 |
| ST485 | 1 | 0.17 |
| ST5266 | 1 | 0.17 |
| ST5281 | 1 | 0.17 |
| ST5468 | 1 | 0.17 |
| ST551 | 1 | 0.17 |
| ST557 | 1 | 0.17 |
| ST564 | 1 | 0.17 |
| ST570 | 1 | 0.17 |
| ST572 | 1 | 0.17 |
| ST6130 | 1 | 0.17 |
| ST622-3LV | 1 | 0.17 |
| ST6274 | 1 | 0.17 |
| ST628 | 1 | 0.17 |
| ST6366 | 1 | 0.17 |
| ST6460-1LV | 1 | 0.17 |
| ST664 | 1 | 0.17 |
| ST6758 | 1 | 0.17 |
| ST70-1LV | 1 | 0.17 |
| ST716 | 1 | 0.17 |
| ST815 | 1 | 0.17 |
| ST895 | 1 | 0.17 |
| ST899 | 1 | 0.17 |
| ST935-1LV | 1 | 0.17 |
| **Resistance score** |  |  |
| 0 | 27 | 4.51 |
| 1 | 230 | 38.4 |
| 2 | 338 | 56.43 |
| 3 | 4 | 0.67 |
| **K locus** |  |  |
| KL15 | 121 | 20.2 |
| KL62 | 63 | 10.52 |
| KL81 | 42 | 7.01 |
| KL102 | 33 | 5.51 |
| KL54 | 19 | 3.17 |
| KL3 | 17 | 2.84 |
| KL25 | 16 | 2.67 |
| KL10 | 15 | 2.5 |
| KL51 | 12 | 2 |
| KL122 | 11 | 1.84 |
| KL19 | 11 | 1.84 |
| KL2 | 11 | 1.84 |
| KL48 | 10 | 1.67 |
| KL114 | 9 | 1.5 |
| KL64 | 9 | 1.5 |
| KL103 | 8 | 1.34 |
| KL17 | 8 | 1.34 |
| KL36 | 8 | 1.34 |
| KL23 | 7 | 1.17 |
| KL8 | 7 | 1.17 |
| KL136 | 6 | 1 |
| KL14 | 6 | 1 |
| KL24 | 6 | 1 |
| KL46 | 6 | 1 |
| KL110 | 5 | 0.83 |
| KL124 | 5 | 0.83 |
| KL125 | 5 | 0.83 |
| KL149 | 5 | 0.83 |
| KL155 | 5 | 0.83 |
| KL105 | 4 | 0.67 |
| KL27 | 4 | 0.67 |
| KL7 | 4 | 0.67 |
| KL1 | 3 | 0.5 |
| KL128 | 3 | 0.5 |
| KL131 | 3 | 0.5 |
| KL132 | 3 | 0.5 |
| KL143 | 3 | 0.5 |
| KL183 | 3 | 0.5 |
| KL20 | 3 | 0.5 |
| KL30 | 3 | 0.5 |
| KL38 | 3 | 0.5 |
| KL52 | 3 | 0.5 |
| KL55 | 3 | 0.5 |
| KL107 | 2 | 0.33 |
| KL109 | 2 | 0.33 |
| KL111 | 2 | 0.33 |
| KL112 | 2 | 0.33 |
| KL123 | 2 | 0.33 |
| KL139 | 2 | 0.33 |
| KL166 | 2 | 0.33 |
| KL167 | 2 | 0.33 |
| KL171 | 2 | 0.33 |
| KL174 | 2 | 0.33 |
| KL186 | 2 | 0.33 |
| KL22 | 2 | 0.33 |
| KL28 | 2 | 0.33 |
| KL47 | 2 | 0.33 |
| KL49 | 2 | 0.33 |
| KL50 | 2 | 0.33 |
| KL56 | 2 | 0.33 |
| KL60 | 2 | 0.33 |
| KL68 | 2 | 0.33 |
| KL106 | 1 | 0.17 |
| KL108 | 1 | 0.17 |
| KL11 | 1 | 0.17 |
| KL113 | 1 | 0.17 |
| KL116 | 1 | 0.17 |
| KL117 | 1 | 0.17 |
| KL119 | 1 | 0.17 |
| KL12 | 1 | 0.17 |
| KL126 | 1 | 0.17 |
| KL13 | 1 | 0.17 |
| KL130 | 1 | 0.17 |
| KL134 | 1 | 0.17 |
| KL135 | 1 | 0.17 |
| KL140 | 1 | 0.17 |
| KL141 | 1 | 0.17 |
| KL144 | 1 | 0.17 |
| KL151 | 1 | 0.17 |
| KL153 | 1 | 0.17 |
| KL157 | 1 | 0.17 |
| KL158 | 1 | 0.17 |
| KL16 | 1 | 0.17 |
| KL179 | 1 | 0.17 |
| KL180 | 1 | 0.17 |
| KL21 | 1 | 0.17 |
| KL34 | 1 | 0.17 |
| KL39 | 1 | 0.17 |
| KL42 | 1 | 0.17 |
| KL5 | 1 | 0.17 |
| KL58 | 1 | 0.17 |
| KL9 | 1 | 0.17 |
| **O locus** |  |  |
| O4 | 158 | 26.38 |
| O1/O2v1 | 128 | 21.37 |
| O1/O2v2 | 128 | 21.37 |
| O3/O3a | 51 | 8.51 |
| OL13 | 47 | 7.85 |
| O5 | 33 | 5.51 |
| O3b | 30 | 5.01 |
| O12 | 12 | 2 |
| OL103 | 9 | 1.5 |
| OL102 | 2 | 0.33 |
| OL104 | 1 | 0.17 |

#### Supplementary Table 2: Summary of genomic features of the 599 KpSC isolates sequenced in this study.

|  |  |  |  | **Unadjusted** | | **Adjusted for Carbapenem resistance** | |
| --- | --- | --- | --- | --- | --- | --- | --- |
| **ST** | Total | Deaths | CFR (%) | OR_CI | p.value | OR_CI | p.value |
| Others | 305 | 106 | 34.8 | - | - | - | - |
| ST11 | 79 | 52 | 65.8 | 3.62 (2.16‚6.16) | <0.001 | 2.41 (1.39‚4.24) | 0.002 |
| ST147 | 34 | 18 | 52.9 | 2.11 (1.03‚4.35) | 0.046 | 1.43 (0.68‚3.03) | 0.343 |
| ST16 | 47 | 27 | 57.4 | 2.53 (1.36‚4.78) | 0.004 | 1.55 (0.80‚3.04) | 0.195 |
| ST1998 | 19 | 11 | 57.9 | 2.58 (1.01‚6.85) | 0.048 | 1.91 (0.73‚5.20) | 0.187 |
| ST48 | 39 | 15 | 38.5 | 1.17 (0.58‚2.31) | 0.648 | 1.19 (0.58‚2.38) | 0.630 |
|  |  |  |  | **Unadjusted** | | **Adjusted for Carbapenem resistance** | |
| **Virulence** | Total | Deaths | CFR | OR_CI | p.value | OR_CI | p.value |
| Others | 578 | 225 | 38.9 | - | - | - | - |
| Hypervirulence | 21 | 4 | 19 | 0.35 (0.10–0.97) | 0.064 | 0.69 (0.19–1.98) | 0.524 |
| Aerobactin | 21 | 4 | 19 | 0.35 (0.10–0.97) | 0.064 | 0.69 (0.19–1.98) | 0.524 |
| Salmochelin | 4 | 1 | 25 | 0.51 (0.03–4.04) | 0.565 | 1.08 (0.05–8.68) | 0.944 |
| RmpA | 21 | 4 | 19 | 0.35 (0.10–0.97) | 0.064 | 0.69 (0.19–1.98) | 0.524 |

#### Supplementary Table 3: Sequence-type (ST) and virulence factor–specific mortality and association with in-hospital death.

### Supplementary Data

#### Supplementary Data 1: Accession numbers and genomic features prediction for 599 KpSC isolates sequenced in the study.
